# The importance of co-located, high intensity smoking cessation support within lung cancer screening: Findings from the Process Evaluation of the Yorkshire Enhanced Stop Smoking study

**DOI:** 10.1101/2024.07.15.24310403

**Authors:** G McCutchan, HD Quinn-Scoggins, H Tong, P Smith, SL Quaife, MEJ Callister, R Thorley, DR Baldwin, RJ Beeken, H Copeland, PAJ Crosbie, S Lewis, S Rogerson, Q Wu, RL Murray, K Brain

## Abstract

**Objective:** Process evaluation of the Yorkshire Enhanced Stop Smoking (YESS) study intervention, to provide evidence regarding optimal integration of smoking cessation support within lung cancer screening (LCS).

**Design:** Mixed-methods process evaluation.

**Setting:** YESS was a Randomised Controlled Trial testing the effect of personalised smoking cessation support, integrated within LCS. YESS study participants were recruited from the Yorkshire Lung Screening Trial.

**Participants/data collection:** Semi-structured interviews with 45 trial participants and eight SCPs 4, 12 and 52-weeks after screening (participants) or training (SCPs). Thematic analysis to assess intervention exposure, context, contamination and theory. Observations of SCP consultations on the screening unit (n=84; 4%) and 4-weeks after screening (n=132; 13%) tested intervention fidelity.

**Intervention:** The YESS study tested opt-out, co-located standard best practice (SBP) smoking cessation support (control) versus a theory-informed personalised risk information booklet designed to increase efficacy beliefs in addition to SBP (booklet intervention), delivered by trained smoking cessation practitioners (SCPs).

**Results:** Intervention context was paramount: participants in both trial arms described benefits of co-located and ongoing high-intensity smoking cessation support, with immediate provision of pharmacotherapy. Tailored, non-judgemental care was considered key to initiating and sustaining quitting, particularly for participants at various points along their quit or those awaiting their scan result. Fidelity was high (98%) and moderate (75%) for SBP, moderate (77%) for the booklet intervention. Exposure varied by participants’ needs, including their screening results. Potential contamination was observed, with SCPs delivering elements of the booklet intervention training across both trial arms.

**Conclusions:** A personalised approach is critical to supporting smoking cessation in LCS. Harnessing the benefits of LCS for supporting cessation at the time of screening requires investment in specialist practitioners to deliver person-centred smoking cessation support.

*Trial registration.* www.clinicaltrials.gov/study/NCT03750110

**Putting research into context:** *What is already known on the topic:*

- Integrated smoking cessation in lung cancer screening is recommended due to the additive benefits of screening participation and cessation on lung cancer mortality.
- Existing evidence supports the provision of higher-intensity smoking cessation interventions within lung cancer screening, such as immediate smoking cessation support at screening, with multiple sessions of behavioural counselling and/or pharmacotherapy.
- However, there is currently no consensus about the optimal high-intensity model to support smoking cessation using behavioural science principles within lung cancer screening. This is a major priority for research, practice and policy.

*What this study adds:*

- We provide evidence for the benefits of co-located and longer-term (up to 12-weeks in person and remote) person-centred support, delivered by trained specialist lung screening SCPs, regardless of trial allocation.
- Future implementation of smoking cessation embedded in lung screening may benefit from investment in specialist lung screening SCPs, adopting a flexible, person-centred approach to the offer and delivery of SBP smoking cessation support.

## INTRODUCTION

Lung cancer screening (LCS) with low-dose computed tomography for high-risk populations based on age and smoking history has been shown to reduce lung cancer mortality,^1,2^ leading to widespread adoption in high-income countries^3^ and recommended implementation in the UK.^4^ Integrated prevention is recommended across LCS programmes due to the additive benefit of screening and cessation on lung cancer mortality (14% additional reduction^5^), and increased cost-effectiveness.^6^ Evidence-informed approaches to integrating smoking cessation and reducing smoking-related inequalities are a major research priority for informing LCS practice and policy.^7^

Various approaches have been tested including low-intensity interventions such as leaflets^8^ and self-/practitioner-referral to external services.^9^ Immediate provision of smoking cessation advice^10^ and higher-intensity interventions such as offering multiple behavioural support sessions and/or pharmacotherapy^10–12^ have shown promise in supporting cessation within LCS. However, high-quality evidence is needed regarding the optimal approach, underpinned by behavioural science principles.^12^ The prevalence of high nicotine dependence and socioeconomic deprivation among lung screening-eligible individuals presents multi-factorial challenges to quitting smoking, reiterating the need for high-intensity approaches in this population.^13^

The Yorkshire Enhanced Stop Smoking (YESS) study^14^ evaluated the effectiveness of an opt-out, co-located standard best practice (SBP) smoking cessation service within the Yorkshire Lung Screening Trial (YLST^15,16^) versus SBP plus a booklet intervention (Figures 1&2). SBP, in line with National Centre for Smoking Cessation standards, consisted of point-of-care behavioural support, pharmacotherapy and 12-weeks of ongoing support with smoking cessation practitioners (SCPs). Given that participation in LCS has been shown to be a key moment to offer cessation support,^17^ research to enhance the potential benefits of SBP further using theory-informed interventions is needed. The booklet intervention, backed by behavioural science, included 12-weeks SBP with an additional personalised booklet intervention incorporating YLST scans of the lungs and heart to indicate any coronary artery calcification and emphysema, or no damage. The booklet, based on the Extended Parallel Processing Model (Figure 2),^18^ was delivered by SCPs following booklet intervention training and using an accompanying script tailored to the extent of heart and/or lung damage. Booklet delivery was designed to further promote increased self-efficacy and response-efficacy beliefs over and above SBP.^19^ Our process evaluation^20,21^ assessed influences on delivery of the YESS booklet intervention and SBP to aid interpretation of the main trial findings (Table 1).

**Figure 1.**
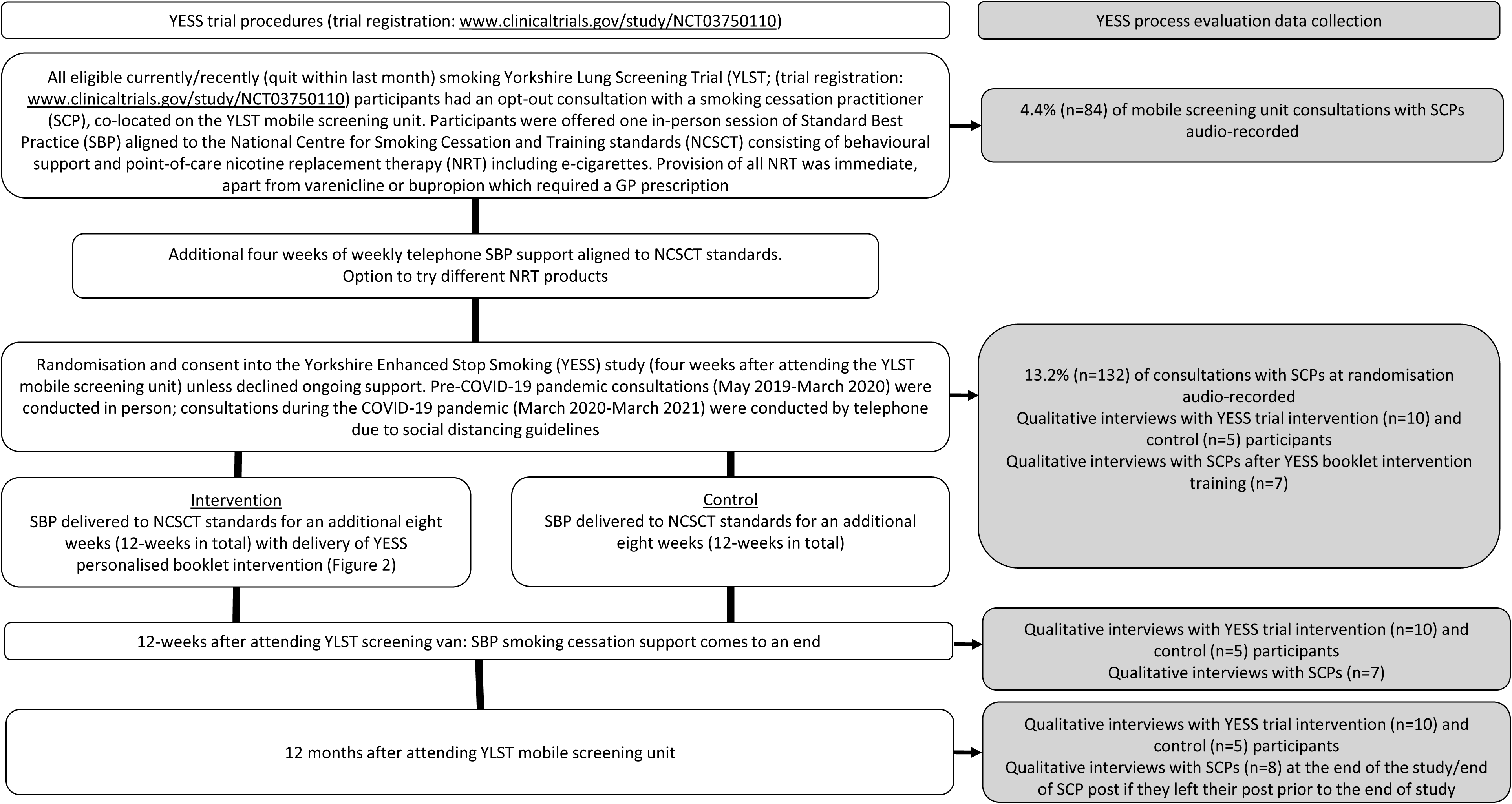
Flow chart of Yorkshire Enhanced Stop Smoking (YESS) study participants and process evaluation components, adapted from Murray *et al* (2020)^14^.

**Figure 2.**
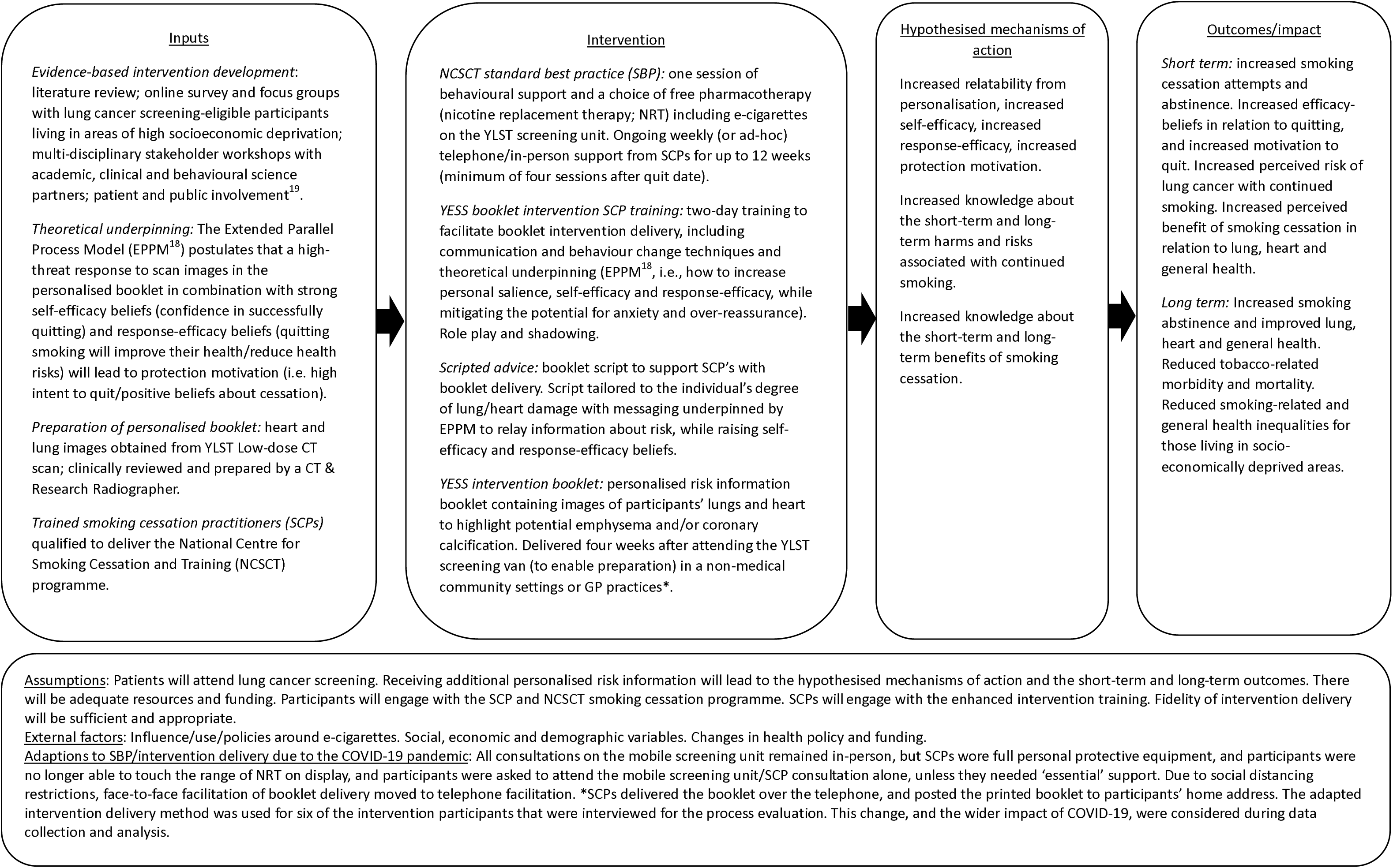
LOGIC model for YESS enhanced.

**Table 1.**
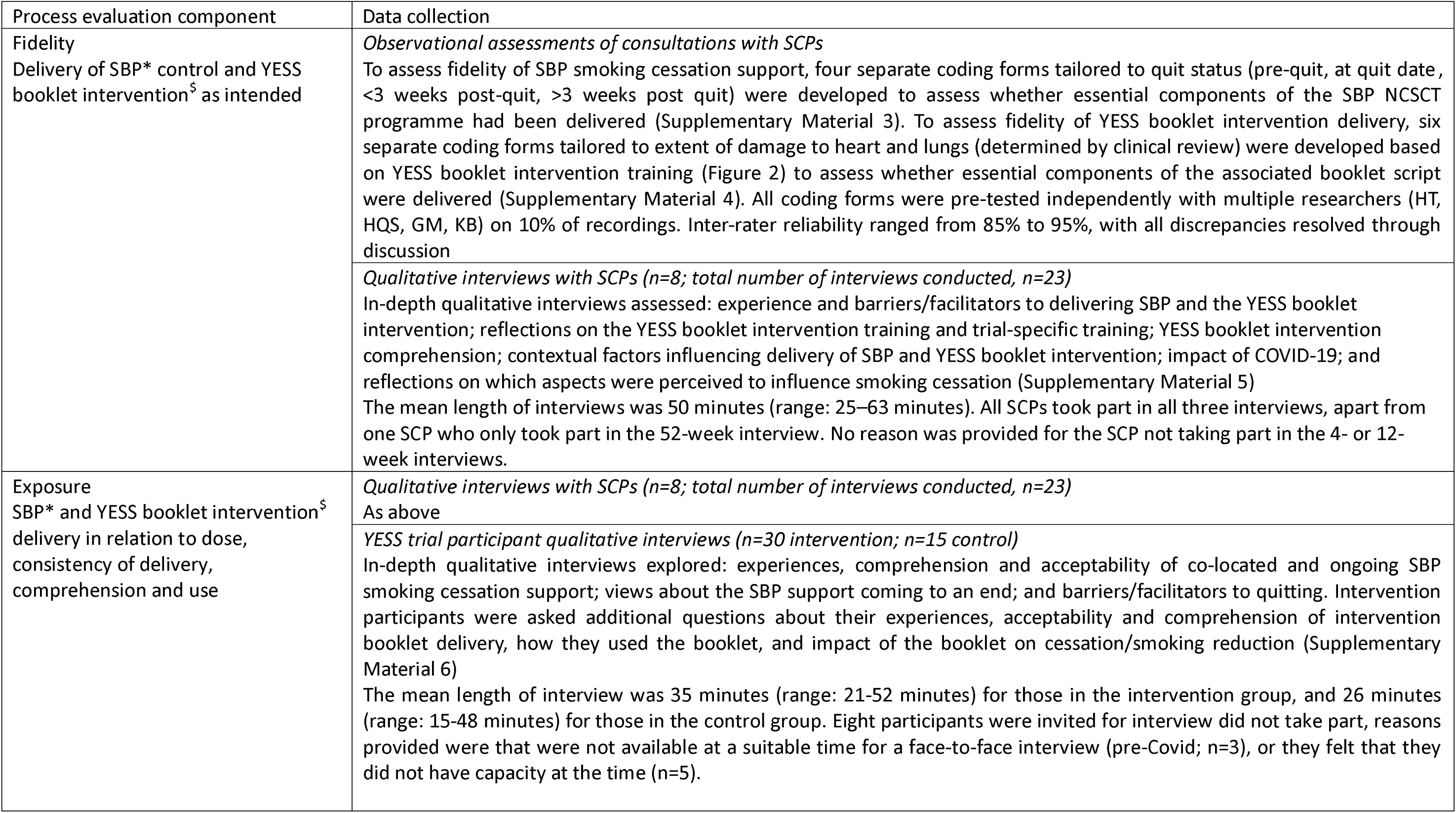
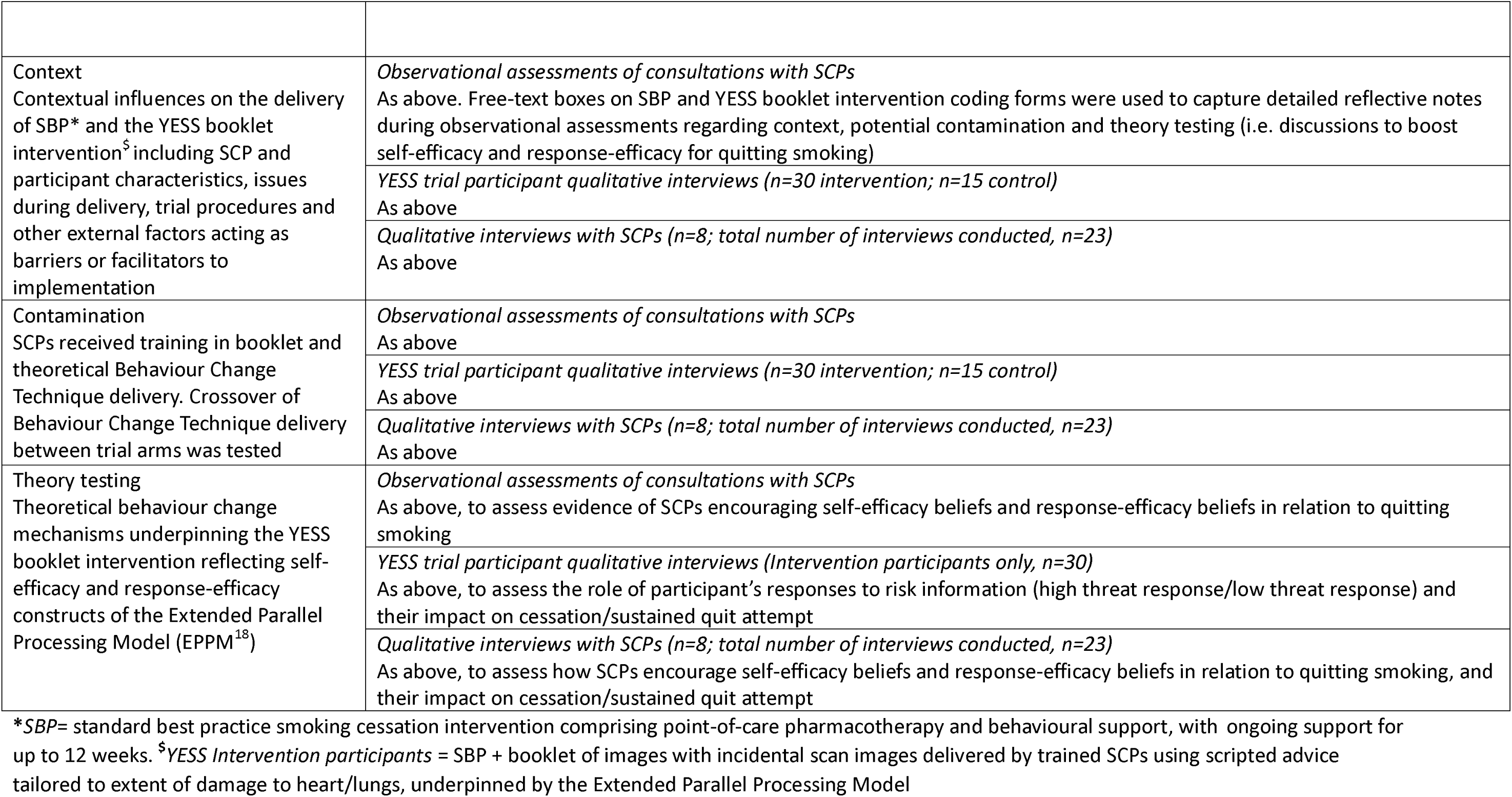
Summary of YESS process evaluation components and data collection methods.

## METHODS

Reporting conforms to STROBE and COREQ guidelines (Supplementary Material 1&2).

### Study design

Mixed-methods trial process evaluation ^21^ to examine fidelity, exposure, contamination, context and theory testing in relation to the main trial findings for the provision of SBP smoking cessation and YESS personalised booklet intervention (Table 1). Data collection took place between April 2019 and April 2021.

### Ethical approval and consent procedures

Trial participants provided consent for observational data and qualitative interviews as part of the YESS consent procedures. ^14^ SCPs provided consent for each interview. Ethical approval was granted by the Health Research Authority, East Midlands-Derby Research Ethics Committee (18/EM/2019).

### Data source and sample

#### Observational data

Audio-recorded consultations with the SCP (i) on the YLST mobile screening unit; and (ii) four weeks after attending YLST screening unit, were sampled purposively based on SCP whole time equivalent, participants’ age and gender, and screening unit location. Incomplete recordings (n=20) were excluded from analysis.

#### Qualitative interviews (YESS participants)

Intervention (n=10) and control (n=5) participants were interviewed at three time points: 4-weeks, 12-weeks and 52-weeks after screening unit attendance (total n=45). Participants were sampled purposively at each time point based on age, quit status and gender, and for intervention participants, extent of heart/lung damage and assigned SCP. Participants were given a £25 shopping voucher.

#### Qualitative interviews (SCPs)

All eight SCPs were invited for sequential interviews at three time points: four, 12 and 52-weeks after the study commenced (or end of employment).

### Data collection

#### Observational data

Audio-recordings were assessed for fidelity of SCPs delivering essential components of SBP and the YESS booklet intervention (Table 1; Supplementary Material 3&4). Separate observational coding forms were used according to quit status (SBP) and booklet (YESS booklet intervention), Table 1. Free-text boxes captured researcher (HT) reflections.

#### Qualitative interviews (YESS trial participants and SCPs)

Semi-structured topic guides were developed to gather in-depth data regarding fidelity, exposure, context, contamination and theory testing (Table 1; Supplementary Material 5&6). Participant interviews were conducted by HQS in-person (n=15) or by telephone due to COVID-19 social distancing restrictions (n=30). SCP interviews were conducted in person (n=12) or by telephone (n=10) due to COVID-19 restrictions.

### Data analysis

#### Observational data

Data were entered into Excel for descriptive analysis. Each essential component delivered scored “1”. Scores were summed to produce total fidelity scores at each time point (SBP delivery: (i) on the screening unit; (ii) 4-weeks; and (iii) YESS intervention booklet delivery at 4-weeks). Due to anticipated variation in the total number of essential components delivered at each time point, by participant quit status, and by intervention booklet type (Supplementary Files 3&4), weighted fidelity scores at each time point are reported. Weighted scores were calculated as the sum of the mean proportion of assessment criteria delivered by quit status/booklet number, multiplied by the number of consultations delivered for that quit status/booklet, and divided by the total number of consultations. Confidence intervals (CI) were calculated at a 95% level. ^22^ Fidelity (percentage of essential components delivered) was categorised as high (>80%), moderate (50%–80%) or low (<50%).^23^

#### Qualitative interviews

Interviews were audio-recorded with permission and transcribed verbatim. Data were analysed thematically^24^ in the stages of familiarisation, development of codes, coding (using NVivo 12; Supplementary Material 7&8) and interpretation of key themes. Codes were initially developed to align with process evaluation components and underpinning theory,^18^ with input from the research team. Ten percent of transcripts were independently dual coded by the research team (HQS, GM or HT), with good agreement, and all discrepancies/amendments to the coding framework resolved through discussion. Two researchers (HQS, GM or HT) independently reviewed all coded data, and met to discuss key themes in a series of three one-day analysis workshops.

#### Data synthesis

Data synthesis workshop including research team members (GM, HQS, HT, PS) to discuss and agree on key findings across data sources that were similar (corroboration), discordant (contradiction), contrasted but generated insights together (complementarity) or enhanced one another (elaboration).^25^

### Patient and Public Involvement (PPI)

Members of the public living in areas of high socioeconomic deprivation and with a smoking history were involved in intervention development. These PPI activities were designed to ensure that the intervention met the needs of the target population.^19^ The Nottingham Tobacco and Nicotine PPI group discussed the main findings of the study, providing their reflections to aid data interpretation. Findings will be disseminated to members of the public via established networks including PPI (e.g. Wales Cancer Research Centre) and via the funder (Yorkshire Cancer Research).

### Role of the funding source

The funders of the study had no role in the study design, data collection, data analysis, or data interpretation, writing of the report, or decision to submit for publication.

## RESULTS

### Sample characteristics

#### Observational data

Fidelity was assessed in 84 (4·4%) complete recordings of SCP consultations on the mobile screening unit. Of SCP consultations at 4-weeks, 132 (13·2%) complete recordings were included. Sample characteristics are shown in Table 2.

**Table 2:** Observational data sample characteristics. *20 incomplete recordings (on screening unit, n=19; four-weeks after attending screening unit, n=1) were removed from analysis due to the recording starting part-way through the consultation or equipment failure meaning the recording was incomplete; N/A: not applicable

|  | Screening unit audio-recorded consultations analysed (n=84*) | 4-week audio-recorded consultations analysed (n=132*) |
| --- | --- | --- |
| *Trial allocation* |  |  |
| Intervention | .. | 68 |
| Control | .. | 64 |
| *Gender identity* |  |  |
| Female | 46 | 65 |
| Male | 38 | 67 |
| *Quit status* |  |  |
| Pre-quit | 83 | 56 |
| At quit date | .. | 4 |
| <3 weeks post quit | 1 | 64 |
| >3 weeks post quit | .. | 8 |
| *COVID-19* |  |  |
| Pre-COVID-19 pandemic (in-person consultation) | 56 | 61 |
| During COVID-19 pandemic (in-person consultation) | 28 | .. |
| During COVID-19 pandemic (telephone consultation) | .. | 71 |
| *Duration of consultation (range in minutes)* | 6m00s - 46m45s | 4m10s – 52m50s |
| *Intervention booklet delivery (range in minutes)* | .. | 4m25s – 19m50s |
| *Booklet type* |  |  |
| No heart damage; moderate lung damage | N/A | 8 |
| No heart damage; no lung damage | N/A | 4 |
| Heart damage; moderate lung damage | N/A | 39 |
| Heart damage; no lung damage | N/A | 16 |
| No heart damage; severe lung damage | N/A | .. |
| Heart damage; severe lung damage | N/A | 1 |

#### YESS trial participant interviews

There was an even split of Intervention and control participants by gender and age. Current smoking was over-represented in SBP participants interviewed. Of the Intervention participants interviewed, most received a booklet showing heart damage and moderate lung damage (Table 3).

**Table 3.** YESS trial participant characteristics (qualitative interviews)

| *Trial allocation* | YESS trial participants (n=45) |  |
| --- | --- | --- |
|  | Intervention (n=30) | Control (n=15) |
| *Gender* |  |  |
| Female | 14 | 9 |
| Male | 16 | 6 |
| *Age: mean years (range)* | 65.2 (57 – 76) | 63.7 (57 – 71) |
| *Index of multiple deprivation* |  |  |
| 1 – most deprived | 10 | 5 |
| 2 | 2 | 2 |
| 3 | 2 | 3 |
| 4 | .. | .. |
| 5 | 3 | .. |
| 6 | 4 | 4 |
| 7 | 3 | .. |
| 8 | 3 | .. |
| 9 | 1 | .. |
| 10 – least deprived | 2 | 1 |
| *Quit status* |  |  |
| Current smoking | 15 | 10 |
| Quit smoking | 15 | 5 |
| *COVID-19* |  |  |
| Interview conducted pre-COVID-19 pandemic | 14 | 8 |
| Interview conducted during COVID-19 pandemic (telephone) | 16 | 7 |
| *Booklet type\** |  |  |
| No heart damage; moderate lung damage | 3 | N/A |
| No heart damage; no lung damage | 2 | N/A |
| Heart damage; moderate lung damage | 18 | N/A |
| Heart damage; no lung damage | 5 | N/A |
| No heart damage; severe lung damage | 1 | N/A |
| Heart damage; severe lung damage | 1 | N/A |
| *no heart damage = no coronary artery calcification; heart damage = presence of coronary artery calcification; no lung damage = no emphysema; moderate lung damage = emphysema but with some preserved areas of normal lung (heterogenous emphysema); severe lung damage = emphysema throughout lung (homogenous emphysema). Heart and/or lung damage determined by clinical review; N/A: not applicable |  |  |

#### SCP interviews

Seven SCPs participated in all three interviews; one SCP participated in the end of study interview. Most SCPs were female (n=6) and had 5-10 years of experience.

### Process evaluation findings

Data from all sources are presented by process evaluation component, with exemplar quotes provided in Table 4.

**Table 4.** Exemplar quotes from qualitative interviews with YESS trial participants and SCPs.

| PROCESS<br>EVALUATION<br>COMPONENT<br>Theme | Data source |  |
| --- | --- | --- |
|  | YESS TRIAL PARTICIPANTS | SMOKING CESSATION PRACTITIONERS (SCPs) |
| FIDELITY |  |  |
| Flexibly delivering SBP | Not applicable | "You've got to be sensitive all the time in this job because you're building that working relationship with people, you're supporting them all the time. The last thing you want to be seen as someone who's not taking on board their concerns or judging them or whatever....I do think we're dealing with a completely different ball game. Yes, the delivery of the content of the stop smoking support is the same because you're working towards the national guidelines. But actually how you deal with the patients is so much different. And it's a way that I have developed through working on the programme. Because as I say when you work in a general stop smoking service, you know, you do have the targets and you... people are coming ready to stop smoking but they're not when they see us here" (*SCP1 12 weeks*) |
| Adapting YESS intervention booklet delivery to SCP's own style and lung cancer screening results | Not applicable | "[I] basically went through the script and picked up all the information I need to deliver...I've just sort of gone about it in my own way...I feel like as long as I'm getting in the key points...If they have got damage, if you stop smoking you can prevent any further damage, or if they haven't got any damage you can stop smoking to prevent it from happening in the future. Obviously go through all the benefits of stopping smoking which are on the back of the leaflet...I do find I just go about in my own way really" (*SCP2, 12 weeks*)<br>"You just get used to working with the patients and dealing with whatever comes up in a sensitive way. Because there's times when |
|  |  | you talk to a patient and they've had a fast-track phone call [for suspected lung cancer]" (SCP1 12 weeks) |
| EXPOSURE |  |  |
| Acceptability and importance of co-located smoking cessation support with a choice of free NRT | *Interviewer:* "How did you feel about being offered to talk to someone about your smoking while you were there [on the mobile screening unit]?"<br>*Participant:* "Absolutely fine, I was more hopeful if anything, that somebody was going to sort of bring the subject up...because I wouldn't have done." (*Intervention, Male, aged 60-64, 4 weeks*)<br>"I couldn't believe it when [SCP] offered me one [e-cigarette]. And she just gave it me. And I could take it off the van that day...Amazing really." (*Control, Female, age 50-54, 12 weeks*) | "I have had people saying that they were lying on the bed [YLST CT scanner] having the screening and they were thinking about what we had just discussed [about smoking cessation]. So that definitely had an impact." (SCP3, 52 weeks)<br>"It's really positive that you are giving them the [support] there and then...they [say] "*Oh I didn't expect this*". It seems entirely right that they should get stuff on the van...The correct way of doing it, as we're doing. To talk to somebody on the van and then say, "*You can go and see [SCP name] in the community in two weeks*" It just seems, I think people just wouldn't go, would they? But us actually being there, I do genuinely think us being on the van is more supportive and conducive to helping people quit, then it would be just referring them to [name of local smoking cessation service]" (SCP4, 4 weeks) |
| Person-centred, non-judgemental and gentle approach to the offer of smoking cessation is vital in a lung screening context | "They don't criticise. They don't start lecturing. They just appreciate what it's like to try and stop. And know full well the best way to help somebody is just to be sympathetic towards it and them and just try and help as much as possible...Yeah totally different [to other services]. I mean not saying they are totally sympathetic, but they weren't critical straight out to you. They will say to you, "*well you do know that it's really advisable that you stop smoking*". And that's about as far as it goes." (*Intervention, Male, aged 70-74, 4 weeks*)<br>"I didn't want to stop, I didn't see the point. It were [SCP name], that convinced me otherwise. [SCP name] said, "*well really why don't you just give it a go, no pressure*." [SCP] tricked me into it [laughs], well no not really tricked me, but just kept going and kept me going, pushing me through. Not in a bad way. After a bit I said | "Reassuring people that we're not going to like tell them off. It's not a case of you've got to do this now... Just easing them into it, letting them work it out at their own pace. I feel like that's definitely needed for the people... Just to reassure them it's completely voluntary, we're not gonna pressure you into doing anything you don't wanna...That way of working is definitely beneficial for these people who weren't necessarily considering stopping smoking until they saw us" (SCP2, 4 weeks)<br>"Some people are a little bit apprehensive because they don't know that they are obviously coming to see you on the van [mobile screening unit]. But once you start having a conversation with them about what the research study provides, the opportunity of not committing to anything for the first four weeks, I find that really really - it's just brilliant. There's no pressure there then. |
|  | *“okay well I’ll just try, no pressure and see”*. And then it weren’t for a few weeks and I realised, well I’ve been going a while now...and one day I was like oh I don’t smoke now!...[It’s] just that [SCP] was there. There for me. Like I didn’t want to stop and he seemed to really understand that. And he didn’t push. We kept going each week... one week at a time...it was week by week and no expectation of the next...I also remember him saying that is was okay, and he understood that it wasn’t easy.” (Control, Female, age 55-59, 52 weeks)<br>“At first I thought here we again, I’m going to get lectured, and they’re gonna talk down to me. But it wasn’t like that. As soon as you walk in they just make you feel at ease straight away...I came back feeling really positive compared to my experience before [group based smoking cessation counselling at GP surgery]. And like I said, I stopped.” (Control, Female, age 60-65, 4 weeks) | Because a lot of these people that you see are really resistant smokers, they’re quite resistant to change. So, giving them that four weeks as well before they commit to anything is just completely different to anything that I’ve done before, it’s brilliant...With [this] research project you get a lot more quality time to spend with the participants to help them... it’s completely different to my previous job...the quality of the service that is provided for the patients is completely different...the follow up appointments and the frequency of them as well, especially in the first four weeks because you contact people once a week for the first four weeks so that’s the time that you allow to be able to, I suppose build up rapport with that person..[in my previous role] usually it was fortnightly appointments.” (SCP5, 4 weeks) |
| Benefits and importance of ongoing support from SCP | “[My SCP] rung every week on a Wednesday... Someone to keep you on the straight and narrow. [My SCP] used to check up on things, and ask if I needed anything else. [SCP] were great. Kept me going.” (Intervention, Female, age 55-59, 12 weeks)<br>“[SCP] spoke to you as a person, not somebody who shouldn’t be smoking. And I absolutely loved that with [SCP]. So I felt more at ease, more comfortable. And I felt that [SCP] has supported me through it for me to stop. And I’ve stopped.” (Control, Female, age 60-65, 4 weeks) | “Generally people do seem to use the fact you are speaking to them on the telephone as a motivator...[Participant] stayed quit because he knew that he would get a phone call from me every week” (SCP1, 4 weeks)<br>“We do say that you’re gonna get tailored support, you’ll be able to speak to me every week, so we’ll be able to keep you on track with how you’re doing. Whereas before most people have never had that sort of support. They’ve just decided one week that they were gonna give it a go, and there’s been no structure to it. But hearing that you’re gonna get 12 weeks, “*hopefully we’ll be able to set a quit date, we’ll keep up with your treatment and I’ll keep up with how you’re getting on. If you’re struggling we’ll cross any bridges when we get them, and we’ll try and cope with anything that’s going wrong*”. And I think hearing that support gives people that bit of motivation to think that this time might be the time that they do actually stop” (SCP2, 52 weeks) |
| Immediate and ongoing SBP with NRT considered to have the greatest impact on quitting compared to the YESS intervention booklet | *Interviewer:* “You had the chat with [name’s SCP] weekly, the [booklet intervention] booklet a few weeks after going to the van, the patches and the vape. Do you think any of it helped more than others?”<br>*Participant:* “I think it all helped...I think the bits [vape and patches] and talking to [names SCP]. Because I knew I should stop. Put it off for years really. But, on the van [mobile screening unit] and with [name SCP] helped me find what would work... [SCP] listened to me and talked through them all [NRT options] and said well I think the vape and the patches would work well.”<br>(*Intervention, Female, age 55-59, 12 weeks*) | “It was the non-judgmental, supportive, on-the-van service that made the difference for me...I think it's more about having the service on the van [mobile screening unit] and the support for 12 weeks rather than the leaflet [intervention booklet]...It's that you get your [NRT] products there and then. You get a friendly person who rings you up once a week and is nice to ya, and is not telling you off....You get the opportunity to try lots of products” (*SCP4, 52 weeks*) |
| EXPOSURE: YESS INTERVENTION BOOKLET |  |  |
| YESS intervention booklet use | “I’ve got [my booklet] here. Had it on my fridge...[It’s] always just up there...I don’t always look though...It’s a reminder for me. Another trick [names SCP] helped me with. If I can’t distract myself [by looking at it] I think well why am I doing this.” (*Intervention, Female, age 70-74, 12 weeks*)<br>*Interviewer:* “And with the booklet... Have you looked it much since, or used it at all?”<br>*Participant:* “No not really...I still have it, I wouldn’t throw it away. I keep all the medical stuff like that. You never know”. (*Intervention, Male, age 55-59, 52 weeks*) | “Maybe something I need to do within my own practice is to remind them to have another look at it, within 12 weeks. So, we're just showing them at four weeks but we could really be reminding them all the way through to refer back to the leaflet. Because I don't know if we really do that enough for the intervention group...We give it them at 4 weeks and then I suppose it's up to them what they do with it. But I think it might be helpful at week eight or so to go get your leaflet out again take a look at it” (*SCP4, 12 weeks*)<br>“What I might do to some people is say “*If you’ve got any cigarettes in the house, them inside the booklet. So you’ve gotta look at that booklet before you have a cigarette*”” (*SCP2, 12 weeks*) |
| YESS booklet intervention comprehension | “[SCP name] was saying at the time it’s good to protect it [healthy areas of heart and lungs]. What you have. By stopping, it will keep it okay.” (*Intervention, Male, 72 years, 52 weeks*)<br>“[SCP said] the stuff about what the cigs were doing to me, and how my health was. Because my heart and lungs are not good, | “It was quite hard to start with... Hard not to sort of over-reassure them on the good images” (*SCP6, 4 weeks*)<br>“Lots of people think they are going to get better, so it’s new to everybody when we tell them they won’t get better” (*SCP4, 4 weeks*) |
|  | especially the lungs...I knew [about the COPD], but this was the first time that anyone had really explained it me...They [the doctors] had told me, and spoken about it, told me what meds I could take and what could happen...But this was the first time someone had really spoken about what it was and why. See I'm not a great one at asking questions see, I like being told what to do and I follow...But with [SCP name] I didn't have to ask questions to get the extra information. [SCP] just went through it all with me and made sure I understood...I've never seen the pictures, the scans before..." (*Intervention, Male, age 60-64, 12 weeks*) | "I have learned a lot as well I think... About emphysema and calcification. I didn't know any of that before. In my previous role it was just a case of helping people to stop smoking, and I knew the implications of carrying on smoking was... But to actually, to deliver these booklets as well... That was quite, that was obviously very new to me... Yeah, I've gained new knowledge. Definitely." (*SCP7, 52 weeks*)<br>"We had one guy who said *"I don't respond well to scaremongering"*. [I'm] having to explain that it's not scaremongering, and it's just a scan of your lung and heart...you get the same kind of sound bites from some people who are in denial and don't want to see that" (*SCP6, 4 weeks*) |
| CONTAMINATION |  |  |
| Potential contamination of YESS booklet intervention components between trial arms | N/A | "I had to be really actually conscious when I was talking to them [control participants]. I had to sort of really think it's a blank piece of paper [indicating control participant], don't tell him anything... I had one woman...She was about 80, but she was really active and independent and getting out and about...It was really difficult, she was really struggling and didn't want to quit those 4 a day...So I had to really talk to her about maintaining her independence...but without relating it to her lung and heart health...It's really hard. You have to be really conscious in what you're saying. And knowing that you've got the standard treatment programme and you've got the intervention... And I'm one of those people that when I've got some information I feel like I've got to share it with everyone. Regardless. So it was really a challenge for me to not share information that I know would benefit her" (*SCP4, 12 weeks*)<br>"[At four-weeks] some people had got their letters back saying that they were, in inverted commas, *"in the clear"*...So [we've] obviously taken that on board as well... Why they should be |
|  |  | quitting smoking and the benefits of it.” (SCP3, 52 weeks) |
| CONTEXT |  |  |
| Lung screening setting | “I guess they have an active audience don’t they? Going for a lung check. And it’s really good. The service...Amazing what they are doing. Doing the scans and all the other tests and then offering to help people stop smoking and providing everything” (Control, Female, age 70-74, 12 weeks)<br>“They’re [YLST staff] just so friendly as soon as you walk in the door. They smile at you and offer you a drink...It makes you feel more comfortable. You’re going to a place you have never been before, and in my mind it was like going to the hospital. At the hospital some staff can be nice and others can be a bit grumpy but every single person on the van, even the girl that took my blood was lovely.” (Control, Female, age 60-65, 4 weeks) | “We’re saying you’re obviously interested in your lung health because you’ve come here today...And what an opportunity this is to make a proper change to protect the health you’ve got now” (SCP4, 4 weeks)<br>“I think coming for the lung health check has sort of sparked, you know, sort of the thought of being at the back of their mind, the thought that they do need to stop smoking” (SCP7, 4 weeks)<br>“I’ve had patients who’ve said that they’ve been thinking about quitting for a long time, and that’s often their wording, and they’ve just not got around to doing anything about it. So coming along to [LHC] and being here, it’s just like perfect ...Sort of good timing – I’ve had a few patients who’ve said that” (SCP1, 4 weeks) |
| Moral obligation to accept smoking cessation support | Interviewer: “What was it then that made you take up the support and carry on then?”<br>Participant: I think I got caught up in it all...it was all a bit much. But that isn’t against [SCP name] or anyone. They were all very kind. And not pushy at all. It was all just a lot. And I didn’t want to say no really while I was there.” (Control, Female, age 65-69, 12 weeks)<br>“Some people, they don’t get that chance do they. To get the help and the checks? We should all feel very lucky.” (Intervention, Female, age 50-54, 12 weeks) | “There are quite a lot of patients who come across to me or wanting to help, because they’re getting a free service so they want to give a bit back... so they feel like because they’re getting something for free then they’re happy to help out with the study, which is how some patients have worded it” (SCP1, 4 weeks)<br>“[one lady] Declined at four weeks, at the four week appointment.... She just said I felt too pressurised when I came the initial visit. She said when I came in, I felt really pressurised. So I said I’m sorry if you felt that way but I assure you it wasn’t my intention. I just wanted to make you aware of what support is available.” (SCP8, 52 weeks) |
| Impact of YLST lung cancer screening results letter | Interviewer: So what one thing made the most difference to you [stopping smoking] then?<br>Participant: Well all of it really...But the letter, the letter from the van saying I had the nodule. That was the last day I picked up and | “I’ve had patients that have had the [YLST results] letter and nodules been on their lung. They’ve had a conversation with you and been open to listening...And realised that this is a warning, this is something that is happening with their health and it’s an |
|  | smoked a cigarette...I thought, no I can't do this anymore. The letter was worded very carefully; I knew it wasn't too awful, but that they would have to do some more tests. So it was scary but it wasn't too, alarming...I threw [the cigarettes] all away. I thought with every cigarette I smoke I am gambling that I will get another one or that this would will be bad. So how could I carry on doing that? (*Intervention, Male, age 65-69, 52 weeks*) | opportunity to be able to change that and turn things around. You just see remarkable changes from patients." (*SCP5, 4 weeks*) |
| Trial-related procedures (CO validation and study questionnaire) used to support conversations about cessation | "It [CO reading] came down from 13 to 3 the last time, so that was an incentive, that was really good. I was made up" (*Intervention, Male, age 60-65, 4 weeks*)<br>"I know a lot of people who have got [COPD]. I know people who have died from cancer. And just...[the high CO reading] just triggers something in your brain....[SCP] said about COPD and that triggered it because I've seen how people have suffered with that. And that's another illness I don't want." (*Control, Female, age 60-64, 4 weeks*) | "We use that questionnaire as a tool because, like the carbon monoxide reading, we're collecting those with the study but we're also using them as a tool to encourage patients to stop smoking. So the questionnaire in a sense is just another tool that we use and it can be a conversation starter." (*SCP8, 12 months*)<br>"I think the carbon monoxide test is a really good way of opening up these conversations with them initially." (*SCP5, 12 months*)<br>"For the vast majority of patients they're really happy when they get a really good non-smoking score [from CO reading]...I think they are a really valuable resource. They help us because it verifies, but it does actually help the patients an awful lot in most cases. Not in every case, but in most cases. They're really delighted when they get a low score." (*SCP1, 12 months*) |
| THEORY TESTING |  |  |
| High threat response | "Well I was scared, yes love. It wasn't nice. It were a shock. Even though I knew something like this was probably coming, it wasn't nice. You know when someone says it. But it didn't traumatise me. It gave me the kick I needed [chuckles]" (*Intervention, Female, age 60-64, 52 weeks*)<br>"How did I feel?...First thing I thought, regarding emphysema, well that's what my Mother died of. And over the last couple of years her breathing was really bad...It were shocking . After seeing that I | "The booklet, I haven't had anyone that's sort of been annoyed in any way. They're a lot more taken aback by the booklet" (*SCP6, 4 weeks*)<br>"The actual, the scan image for people who are on the intervention, I feel like that is a really good bit of information for them to have. Everyone knows the certain statistics around smoking, like X number of people that smoke will get this disease...And a lot of people think well if it happens so be it, or |
|  | thought I've got to do something. I thought, [the SCP on the mobile screening unit] were talking sense. And then I thought I've got to stop. But then when I saw them images [booklet] I thought all the more then. I'll be stupid if I don't." (*Intervention, Male, age 70-75, 4 weeks*)<br>"It were, not nice shall we say....[I thought] This is my own doing this. And so when I saw [the intervention booklet], well that was it. I couldn't do it [smoke] no more...Every time I had one. It felt dirty. Like I could feel it in my lungs and chest. Like I hadn't before" (*Intervention, Male, age 60-64, 12 weeks*) | they'll think it's not gonna happen to me so don't matter...But with the actual images seeing the damage and what they are doing, their own body, that can be a big sign...I'm not invincible, I am actually damaging myself here, I am killing myself, I need to do something about it....It works both ways... So alot of damage here, but on the flip side if their can results are not showing anything they could think "*Oh well I'm alright, I can carry on doing it [smoking] then*" ... So you have to say it could happen at any time" (*SCP2, 4 weeks*) |
| Building self-efficacy and response efficacy beliefs in relation to intervention booklet images | *Participant:* "We had a long talk about it. And [SCP] was saying that you know I'm okay on the outside. But that it could get bad...It [emphysema] can always get bad or make you ill at any point."<br>*Interviewer:* "Had you thought about it like that before?"<br>*Participant:* "Well I guess not. I tend to be one for living in the moment... And [names SCP] they made me realise that the moments won't always be there if I get sick. Or if my lungs start packing in." (*Intervention, Female, age 60-64, 12 weeks*) | "I don't want them to cry or anything but I do want them to realise the seriousness, that it's serious... So it's kind of finding the balance between, you know, this is damage and has been caused by smoking, especially the lung images, and the best thing you can do is to stop smoking... Make sure they understand and relate to the images and that they use the images going forward to remind themselves" (*SCP1, 4 weeks*)<br>"I think the leaflet helps...It helps to focus and it's a good tool to help people think about their own smoking and the consequences of smoking" (*SCP4, 12 weeks*) |
| Protection/defensive motivation | "[seeing the booklet], I were a bit gutted actually to tell you the truth...[names SCP] explained everything to me, what this and what were. And I thought, right that's it, no more cigs for me... I want to live to see my grandkids grow up." (*Intervention, Female, age 75-79, 4 weeks*)<br>"I had mild emphysema... I walked out of there [4 week consultation] and I thought that's it, you've got to do it [quit smoking]. If you do anything you're just going to be like your Dad [who died of COPD], and I don't want my kids or my grandkids to see me like that...My kids wouldn't go see my Dad because he were | "I think that shocked and upset feeling has been the driving force in making them quit smoking. Or not making them quit smoking, but then deciding that they need to quit smoking" (*SCP1, 52 weeks*)<br>"[one participant] struggled, she didn't want to see the leaflet...I sent it to her but then I rang her later and she said she had opened it but then put it straight in the bin..." (*SCP4, 52 weeks*) |
|  | laid in hospital and he had a full face mask on because he couldn't breathe" (*Intervention, Male, age 55-59, 4 weeks*) |  |

**Footnote:** Quotes are presented in smart verbatim; '...' denotes text removed. Text in square brackets [] denotes text inserted by the authors for context, or where identifiable information has been concealed. Text in round brackets () refers to participant characteristics and time point of data collection. SCP gender and age are concealed due to being potentially identifiable.

### Fidelity

SBP was delivered with high fidelity (98·0%, CI:96·6% to 98·9%) on the screening unit, and with moderate fidelity (74·6%, CI:72·1% to 76·6%) four weeks after attending the screening unit, Table 5. SCPs described prioritising aspects of SBP based on participant needs or time constraints.

**Table 5.**
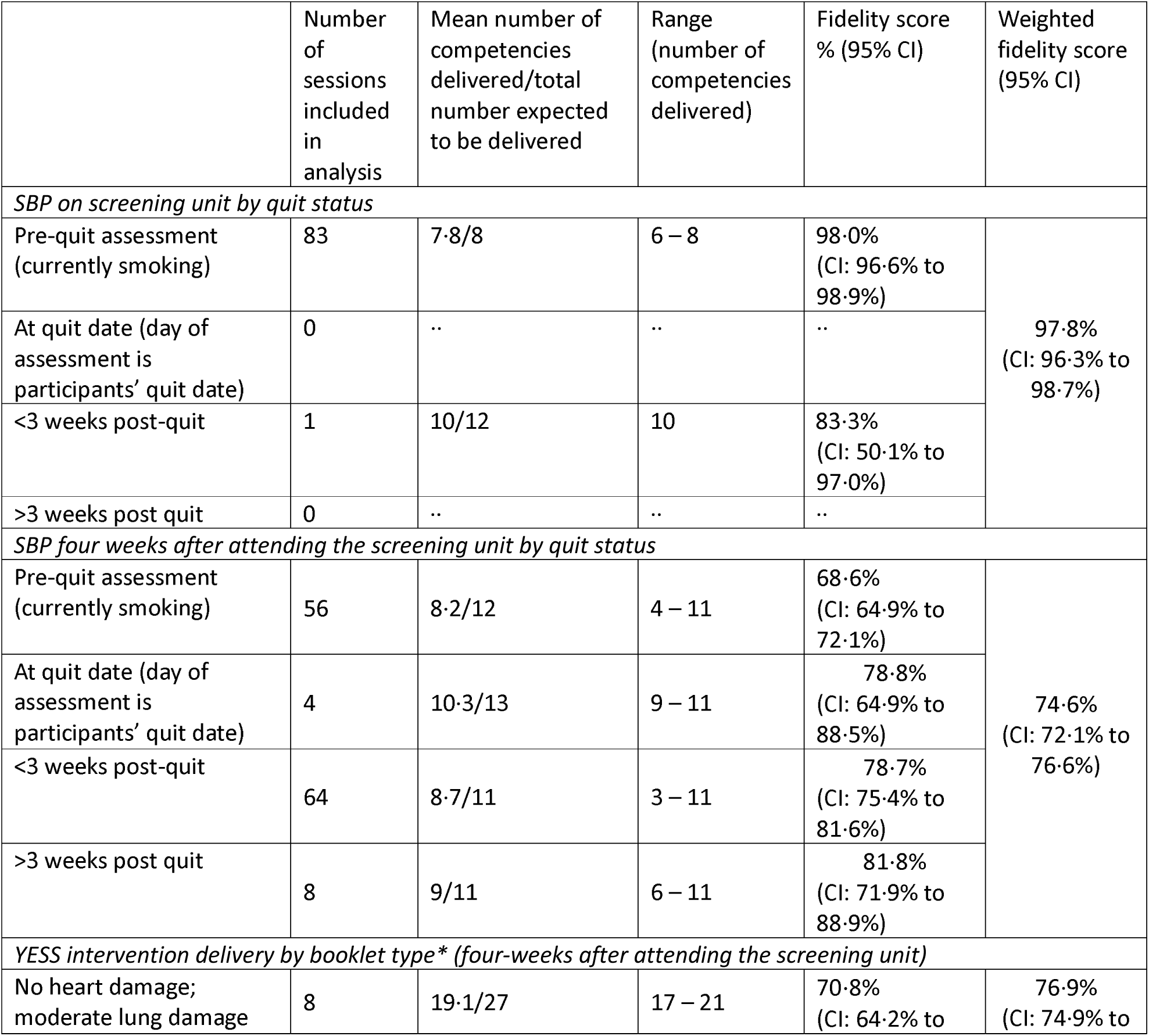
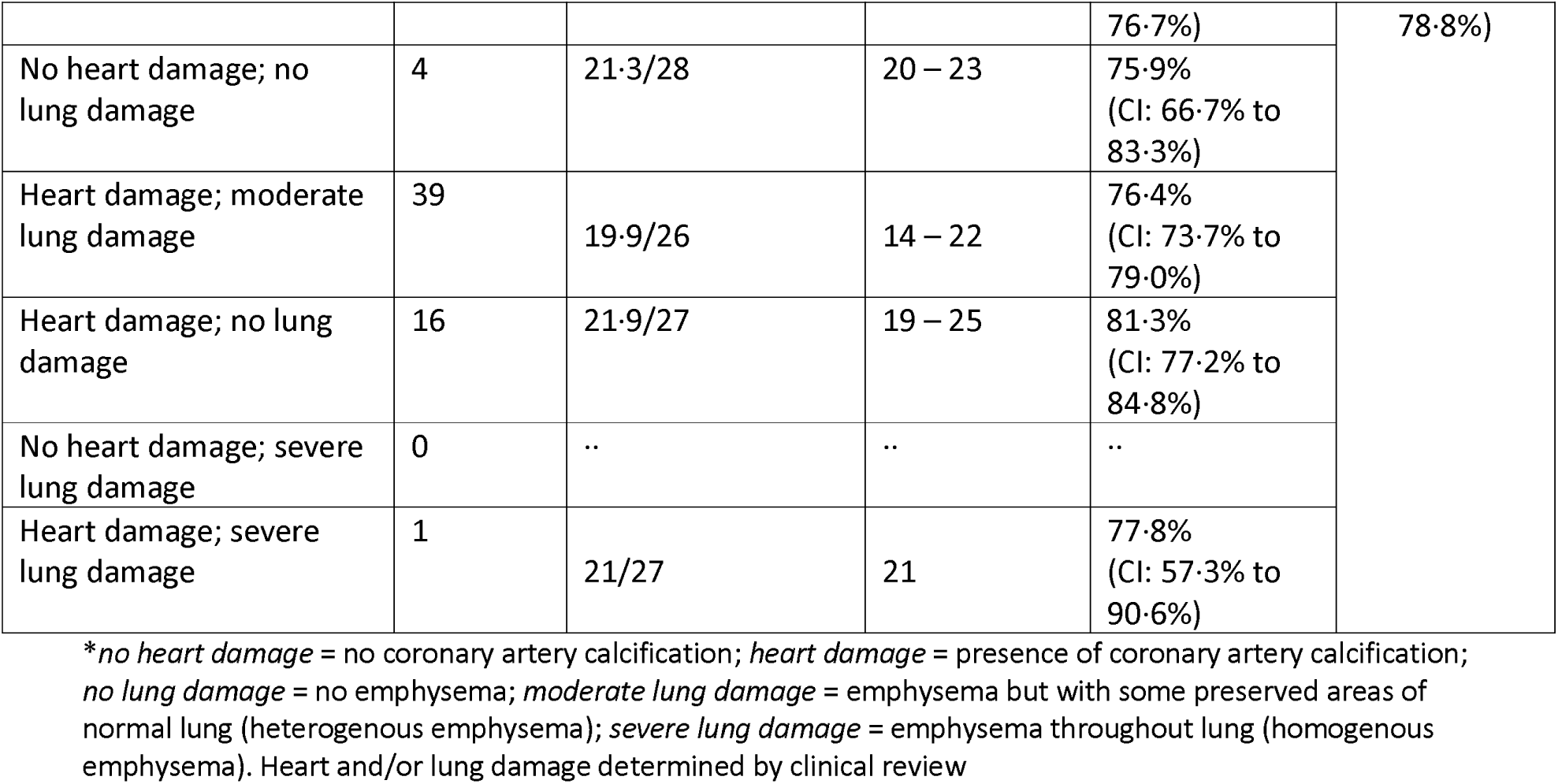
Fidelity of SBP and YESS booklet intervention.

The YESS intervention booklet was delivered with moderate fidelity (76·9%, CI:74·9% to 78·8%), Table 6. All SCPs understood the importance of delivering the intervention booklet consistently, but also discussed adopting their own style of delivery in practice. Some SCPs tended to focus on what they considered “key points”, while other SCPs relied heavily on the script. When participants had been urgently referred on lung cancer diagnostic pathway (“fast-tracked”) or diagnosed with lung cancer, SCPs described approaching booklet delivery sensitively and some SCPs offered the option to opt-out or to revisit the booklet at a later date.

### Exposure: SBP

#### SBP delivery: behavioural support and pharmacotherapy

All participants described the acceptability and importance of having SCPs co-located on the mobile lung screening unit. SCPs felt that the combination of high-intensity, immediate and ongoing SBP support with a range of point-of-care, free nicotine replacement therapy (NRT), e-cigarettes aided or maintained abstinence, and was considered to have greatest impact on quitting, irrespective of trial arm. SCPs described LCS being a unique context and cohort of patients with varying levels of quit motivation, anxiety about screening, and often with complex social and health needs. A person-centred, flexible approach to the offer of smoking cessation support was considered vital. In both trial arms, SCPs described tailoring discussions about barriers to quitting and the benefits of cessation based on the individual’s quit motivation and emotional state. For those not ready to set an immediate quit date, SCPs described using the first four weeks (prior to randomisation), as a buffer period to increase participants’ interest and confidence in trying NRT. All trial participants described a trusting relationship with their SCP, founded on non-judgemental and compassionate care, and participants felt valued and worthy of the resource invested in them.

#### Quality and quantity of SBP

SCPs considered that referral to external stop-smoking services would not have the same impact on quit rates. They compared their experience working on YESS with their personal experience working with external smoking cessation services. Compared to their previous roles, SCPs described higher quality SBP with greater flexibility afforded within YESS, including absence of quit rate targets, more intensive support (e.g. weekly/longer appointment times), a broader range of NRT and provision of e-cigarettes. These adjustments were considered to positively impact on participants’ receptivity and motivation to quit. Co-location on the mobile unit, with a low pressure commitment, flexible appointment times to fit around participants’ work, and in-person/remote support to confidently use NRT e.g. share videos, reduced access barriers for participants across both trial arms.

### Exposure: YESS booklet intervention

#### YESS booklet intervention delivery

SCPs initially felt apprehensive about delivering the booklet to intervention participants who may become upset. However, SCP training and the associated booklet script boosted SCP confidence to deliver the booklet intervention. SCPs found delivering booklets with extensive heart and/or lung damage challenging and “draining”, although easier (“less of a blow”) to deliver when Intervention participants disclosed a prior diagnosis of co-morbid heart and/or lung condition(s).

#### YESS booklet intervention use

Intervention participants were generally receptive to, and grateful for the booklet. All 4-week and 12-week, and most 52-week intervention participants kept their booklet, usually somewhere safe e.g., with other medical information. Most had shown their booklet to someone close to them, e.g. a spouse or trusted friend. However, some with extensive heart and/or lung damage preferred not to share their diagnosis with family or friends to avoid worrying them. A few participants (mostly women) shared their booklet more widely i.e. to friends/family via WhatsApp, and some visited their GP to discuss the booklet. There were differences in how participants used their booklet. Some, mostly female participants, regularly referred back to their booklet to remain abstinent e.g. displaying it on the fridge/on top of cigarette packets, while others did not refer back to the booklet. Differences in booklet use may reflect variation in SCP delivery: some SCPs actively advised participants to refer back to their booklet/place it somewhere salient; other SCPs tended not to mention the booklet again.

#### Comprehension

SCPs ensured intervention participants understood that the images in the booklet were their own and that damage could not be reversed, while trying to strike a balance between over- and under-reassurance of results. All intervention participants demonstrated good levels of comprehension that stopping smoking could protect their health by preventing (further) calcification/emphysema, and that existing damage could not be reversed. However, some misinterpreted the images i.e. assumed the lung not shown was healthy. For intervention participants with previous known diagnoses of lung and/or heart problems, most described truly understanding the damage for the first time because the SCP explained their diagnosis in lay language with the scan images. SCPs recounted a few consultations where intervention participants were dismissive of the booklet: some queried whether the images were their own, or whether the damage was smoking-related.

### Contamination

#### Potential contamination of YESS intervention components between trial arms

SCPs enjoyed the theory and lung health aspects of the intervention booklet training, and felt elements of the bespoke training improved their practice. One SCP described being actively conscious of potential crossover of booklet intervention components to the control arm, as per their training. However, there was evidence suggestive of contamination: that SCPs drew on elements of the booklet intervention training with participants across both trial arms. For example, when SCPs used participants’ attendance at lung screening to steer conversations about the additive benefits of quitting to further protect lung health (see *Context* section), SCPs used similar phrasing from the intervention booklet script/training. Further, although not part of the booklet intervention or SBP, participants tended to discuss their LCS results with the SCPs. By the 4-week SCP appointment, most participants had received their LCS results letter (all clear/further investigations/”fast-tracked” for suspected lung cancer). SCPs tended to utilise the screening result to encourage efficacy beliefs initiating or sustaining a quit attempt, drawing on the health-benefits of cessation, and tailoring discussions to their screening result e.g. SCPs approached conversations with fast-tracked participants with extreme sensitivity, but reiterated the benefits of quitting for their health, any planned treatment and prognosis.

### Context

#### Lung cancer screening setting

Trial participants across both arms described welcoming, friendly and compassionate staff throughout the YLST mobile screening unit. SCPs felt this positively impacted on screening attendees being more receptive to the offer of smoking cessation support. However, some participants across trial arms (particularly women) described feeling a moral obligation to accept support. Being invited to a lung health service set-up specifically to help people with a long-term history of smoking enabled participants across both trial arms feel valued. The lung screening context, in which participants had pro-actively engaged in a service for their lung health, was used by SCPs to steer conversations around screening as an opportunity to support engagement with the service for participants in both trial arms. For most trial participants in both arms, the YLST LCS results letter (regardless of result) acted as an additional motivator to consider making a change to their smoking, and SCPs tailored SBP support in both trial arms to LCS result. Intervention participants tended to place more value on their YLST letter versus booklet.

#### Trial-related procedures

Elements of the trial questionnaire, which was completed by SCPs, acted as an additional tool to support cessation conversations e.g., to identify prior NRT use/barriers to quitting, enabling SCPs to then build participants’ response-efficacy beliefs in relation to quitting. For example, with participants who indicated worry about lung cancer on the questionnaire, SCPs would focus discussions on the benefits of quitting to protect lung health and reduce their risk of lung cancer. Carbon Monoxide (CO) readings obtained for the trial (although can be utilised in SBP) acted as a personalised intervention, with high readings eliciting a high-threat response. Seeing CO readings decrease to mirror “non-smokers” provided motivation and immediate external validation.

### Theory testing

#### Threat response

All intervention participants described an emotive, high-threat response to the booklet, regardless of levels of damage. Response was intensified for people with emphysema in their family and/or social network. Vitally, SCPs managed emotional reactions to the booklet. No intervention participants described an initial low/no threat response to the booklet, although threat responses were lower by the 52-week interviews. Some intervention participants described feeling guilt and shame when smoking after seeing the booklet images, and pictured imagery of dirt entering their lungs when smoking again.

#### Response-efficacy (belief that stopping smoking would benefit/protect their health)

All intervention participants exhibited strong response-efficacy beliefs in response to their booklet. SCPs described using the booklet as another excellent tool (in addition to CO validation, trial questionnaire, NRT products and screening results letter) to support discussions about the consequences of smoking, particularly in the context of LCS. Participants felt the personalisation of the booklet containing their own scan images was of greater impact than images on cigarette packets or generic health information.

#### Self-efficacy

Discussions with SCPs around the intervention booklet further reinforced participants’ reported beliefs about confidence in quitting that had been encouraged in earlier discussions as part of SBP.

#### Protection motivation/defensive motivation

Most intervention participants had quit smoking or decided to quit prior to receiving the booklet, and the booklet helped them to sustain abstinence. SCPs felt that the booklet was most impactful for intervention participants who were considering, but unsure about quitting (i.e. “on the fence”), because they felt empowered and motivated to quit. For a small minority of intervention participants who responded to their booklet with strong emotional reactions/rejection, this led to disengagement with the booklet and service.

## DISCUSSION

This was the first study to highlight the importance of providing immediate behavioural smoking cessation support with pharmacotherapy, physically co-located within LCS. High-intensity and long-term SBP smoking cessation support using a person-centred approach was considered vital to boost receptivity and motivation to quit, and to maintain abstinence in both trial arms. SCPs delivered the booklet intervention flexibly by adapting delivery of the booklet intervention to individual need. SCPs described variably in their approach for how they deliver the booklet. There was evidence of possible contamination of the booklet intervention components between trial arms. In line with the SCPs booklet intervention training, the LCS context (attendance at screening and results letter) was used by SCPs in both trial arms to boost response-efficacy beliefs about the benefits of quitting. Participants felt that reliable, long-term SBP had the greatest impact in supporting smoking cessation in a LCS context, over-and-above the booklet intervention.

We provide further evidence for the benefits of immediate^10^ and high-intensity^11^ smoking cessation support within LCS. Our process evaluation extends these findings to emphasise the importance of co-located services, with longer-term (up to 12 weeks) person-centred and non-judgemental support, for engaging individuals with high nicotine dependence in smoking cessation support^26^. The lack of intervention effect on quit rates observed in YESS (Murray et al, submitted^27^) could potentially be explained by the high standard of SBP delivered four-weeks prior to, and in conjunction with the booklet intervention; in turn potentially limiting the ability of the booklet intervention to provide additive benefit. Potential contamination of the booklet intervention between trial arms could also explain the lack of intervention effect. Previous research has shown that participation in LCS and receiving LCS results can increase motivation to quit smoking.^28^ In this study, SCPs across both trial arms used participants’ attendance at LCS and/or LCS results to highlight the added benefit of cessation for their lung health. This was influenced by the training received as part of the booklet intervention and highlights the potential value of a specialist lung screening SCP model, similar to specialist models used in other contexts e.g. pregnancy.^29,30^

Our robust process evaluation is the first to provide rich insights regarding the importance of high-intensity, co-located smoking cessation support within LCS. We rigorously collected and analysed data across multiple timepoints and data sources, adding important evidence from the perspective of both SCPs and service-users in relation to integrated smoking cessation in LCS. Limitations of the study include possible sampling/selection bias for the observational assessments, in which SCPs selected consultations to record and resulted in some SCPs recording more consultations than others. Further, due to the process evaluation design, we were unable to explore gender effects of the booklet intervention in detail in relation to main trial findings, or confirm contamination between trial arms due to subtle nuances of communication, and this warrants further research. Future controlled evaluations of smoking cessation interventions in lung screening could address issues of contamination by having different SCPs deliver the control and intervention arms.

Point-of-care/ongoing NRT provision combined with behavioural support within the screening context (i.e. attendance and results letter) can be used positively to encourage quitting. Future implementation of smoking cessation embedded in lung screening may benefit from investment in specialist ‘lung screening-experienced’ SCPs,^29,30^ with enhanced SBP training to further develop SCPs’ skills in building self-efficacy and response-efficacy beliefs in the context of LCS. Specialist lung screening SCPs could adopt a flexible, person-centred approach to the offer and delivery of SBP cessation support, enabling tailored support based on quit motivation, the lung screening context and wider determinants of health in the high-risk population typically experiencing long-term tobacco dependence.^13^ A buffer period between screening attendance and quit date can be used to increase participants’ interest and confidence in using NRT prior to abrupt cessation, particularly for participants not considering quitting at the time of screening. However, maintaining, delivering and monitoring the high standards of SBP delivered within this study may be challenging in a real-world setting.

This study provides novel, high-quality evidence for the benefits of integrated and ongoing SBP smoking cessation support within LCS. Due to the unique context and cohort of lung screening patients, support for quitting smoking should be delivered flexibly by specialist SCPs.

## Data Availability

Data sharing of anonymised data is available upon reasonable request. Please contact the corresponding author for a data-sharing request form. Data-sharing requests will be considered for a methodologically sound proposal. The request should detail clear objectives, what data are requested, timelines for use, intellectual property and publication rights, data release definition in the contract and participant informed consent, etc. A data-sharing agreement from the sponsor may be required.

## Role of the funding source

The funders of the study had no role in study design, data collection, data analysis, data interpretation or writing of the report.

## Declaration of interests

RT, PC have received payment/honoraria for a webinar with Bayer. DRB has received lecture honoraria with Astra Zeneca, MSD and Roche. RM has received consulting fees from Action on Smoking and Health and Cancer Research UK. RM is a Trustee of Action on Smoking and Health and a member of the Royal College of Physicians Tobacco Advisory Group. All other authors declare no conflicts of interest.

## Conflicts of interest

Interests declared as above. No other conflicts of interest.

## Acknowledgements

We thank all participants who took part in the study. We acknowledge the contribution of all the YESS study team (Gail Barden, Jannine Brooks, Kate Fraser, Thomas Griffiths, Matthew Hudson, Sue O’Shea, Dianne Preesha, Susan Tyrrell, Jill Young, Rebecca Thorley, Harriet Copeland, Alex Ashurst) and the whole YLST teams in delivering these interventions. This work was funded by Yorkshire Cancer Research (award references L403 and NOT414). Thanks to Prof Daniel Farewell for his advice on calculating fidelity scores. From September 2021, P. Alexandris was supported by the Barts Charity (MRC&U0036). From January 2021, S.L. Quaife was supported by the Barts Charity (MRC&U0036). R.J. Beeken is supported by a Yorkshire Cancer Research Fellowship (L389RB). P.A.J. Crosbie is supported by the Manchester National Institute for Health Research Manchester Biomedical Research Centre (IS-BRC-1215-20007). H. D. Quinn-Scoggins is supported by Health and Care Research Wales as part of the Primary and Emergency Care Research Centre (PRIME) (517195). G. McCutchan is supported by Health and Care Research Wales as part of the Wales Cancer Research Centre (517190).

## Author contributions

RM and MC co-led the main YESS trial. The YESS process evaluation was designed by KB, with substantial input and expert advice from HQS, GM, RM, SQ and RT, and the YESS TMG. Qualitative process evaluation data was collected by HSQ. Observational process evaluation data collection was co-ordinated by HSQ and RT (audio-recordings taken by SCPs during consultations). Bespoke observational data coding forms were initially developed by HSQ, with expert input from KB, RM, SQ, RT, GM and HT. HSQ, GM, HT and PS conducted process evaluation data analysis, overseen by KB and RM. GM drafted the manuscript, and all authors contributed to the review and editing of the manuscript. All authors read and approved the final manuscript. The corresponding author attests that all listed authors meet authorship criteria and that no others meeting the criteria have been omitted.

## Transparency statement

The lead author affirms that the manuscript is an honest, accurate, and transparent account of the study being reported. No important aspects of the study have been omitted.

## Copyright/license for publication

The Corresponding Author has the right to grant on behalf of all authors and does grant on behalf of all authors, a worldwide licence to the Publishers and its licensees in perpetuity, in all forms, formats and media (whether known now or created in the future), to i) publish, reproduce, distribute, display and store the Contribution, ii) translate the Contribution into other languages, create adaptations, reprints, include within collections and create summaries, extracts and/or, abstracts of the Contribution, iii) create any other derivative work(s) based on the Contribution, iv) to exploit all subsidiary rights in the Contribution, v) the inclusion of electronic links from the Contribution to third party material where-ever it may be located; and, vi) licence any third party to do any or all of the above.

## Notes

### Clinical Trial

NCT03750110

### Clinical Protocols

https://doi.org/10.1136/bmjopen-2020-037086

### Funding Statement

This work was funded by Yorkshire Cancer Research (award references L403 and NOT414). From September 2021, P. Alexandris was supported by the Barts Charity (MRC&U0036). From January 2021, S.L. Quaife was supported by the Barts Charity (MRC&U0036). R.J. Beeken is supported by a Yorkshire Cancer Research Fellowship (L389RB). P.A.J. Crosbie is supported by the Manchester National Institute for Health Research Manchester Biomedical Research Centre (IS-BRC-1215-20007). H. D. Quinn-Scoggins is supported by Health and Care Research Wales as part of the Primary and Emergency Care Research Centre (PRIME) (517195). G. McCutchan is supported by Health and Care Research Wales as part of the Wales Cancer Research Centre (517190).

### Author Declarations

Ethical approval was granted by the Health Research Authority, East Midlands-Derby Research Ethics Committee (18/EM/2019).

