## Supplementary material for "The importance of co-located, high intensity smoking cessation support within lung cancer screening: Findings from the Process Evaluation of the Yorkshire Enhanced Stop Smoking study": suppl File

### Table of Contents


| Supplementary material # | Title | Page numbers |
|---|---|---|
| 1 | STROBE Statement checklist of items that should be included in reports of observational studies | 2-4 |
| 2 | Consolidated criteria for reporting qualitative studies (COREQ): 32-item checklist with page numbers to indicate section of the article | 5-6 |
| 3 | Assessment criteria for the delivery of essential components of standard best practice by quit status on the screening van and at four-week assessment[1] | 7 |
| 4 | Assessment criteria for the delivery of essential components of the YESS Booklet intervention by booklet number at four-weeks[1] | 8-9 |
| 5 | Topic guides for interviews with SCPs | 10-13 |
| 6 | Topic guides for interviews with trial participants | 14-24 |
| 7 | SCP interview coding framework (combined across timepoints) | 24-28 |
| 8 | trial participant interview coding framework (combined across timepoints) | 29-33 |

### Supplementary Material 1. STROBE Statement checklist of items that should be included in reports of observational studies

| | Item No | Recommendation | Page number in article |
|---|---|---|---|
| **Title and abstract** | 1 | (*a*) Indicate the study's design with a commonly used term in the title or the abstract | 1 and 4 |
| | | (*b*) Provide in the abstract an informative and balanced summary of what was done and what was found | 4 |
| **Introduction** | | | |
| Background/rationale | 2 | Explain the scientific background and rationale for the investigation being reported | 5 |
| Objectives | 3 | State specific objectives, including any prespecified hypotheses | 5 |
| **Methods** | | | |
| Study design | 4 | Present key elements of study design early in the paper | 8 |
| Setting | 5 | Describe the setting, locations, and relevant dates, including periods of recruitment, exposure, follow-up, and data collection | 8 and Table 1 |
| Participants | 6 | (*a*) *Cohort study*—Give the eligibility criteria, and the sources and methods of selection of participants. Describe methods of follow-up<br>*Case-control study*—Give the eligibility criteria, and the sources and methods of case ascertainment and control selection. Give the rationale for the choice of cases and controls<br>*Cross-sectional study*—Give the eligibility criteria, and the sources and methods of selection of participants | 8 and Table 1 |
| | | (*b*) *Cohort study*—For matched studies, give matching criteria and number of exposed and unexposed<br>*Case-control study*—For matched studies, give matching criteria and the number of controls per case | N/A |
| Variables | 7 | Clearly define all outcomes, exposures, predictors, potential confounders, and effect modifiers. Give diagnostic criteria, if applicable | Table 1 |
| Data sources/ measurement | 8* | For each variable of interest, give sources of data and details of methods of assessment (measurement). Describe comparability of assessment methods if there is more than one group | 8 and Table 1 |
| Bias | 9 | Describe any efforts to address potential sources of bias | N/A |
| Study size | 10 | Explain how the study size was arrived at | 8 |

| | | | |
|---|---|---|---|
| Quantitative variables | 11 | Explain how quantitative variables were handled in the analyses. If applicable, describe which groupings were chosen and why | 8-9 |
| Statistical methods | 12 | (*a*) Describe all statistical methods, including those used to control for confounding | 9 |
| | | (*b*) Describe any methods used to examine subgroups and interactions | 9 |
| | | (*c*) Explain how missing data were addressed | 9 and Table 1 |
| | | (*d*) *Cohort study*—If applicable, explain how loss to follow-up was addressed<br>*Case-control study*—If applicable, explain how matching of cases and controls was addressed<br>*Cross-sectional study*—If applicable, describe analytical methods taking account of sampling strategy | Table 1 |
| | | (*e*) Describe any sensitivity analyses | N/A |

**Results**

| | | | |
|---|---|---|---|
| Participants | 13* | (a) Report numbers of individuals at each stage of study—eg numbers potentially eligible, examined for eligibility, confirmed eligible, included in the study, completing follow-up, and analysed | 9 and Table 2 |
| | | (b) Give reasons for non-participation at each stage | - |
| | | (c) Consider use of a flow diagram | - |
| Descriptive data | 14* | (a) Give characteristics of study participants (eg demographic, clinical, social) and information on exposures and potential confounders | 9 and Table 2 |
| | | (b) Indicate number of participants with missing data for each variable of interest | 10 |
| | | (c) *Cohort study*—Summarise follow-up time (eg, average and total amount) | N/A |
| Outcome data | 15* | *Cohort study*—Report numbers of outcome events or summary measures over time | |
| | | *Case-control study*—Report numbers in each exposure category, or summary measures of exposure | |
| | | *Cross-sectional study*—Report numbers of outcome events or summary measures | 11-13 |
| Main results | 16 | (*a*) Give unadjusted estimates and, if applicable, confounder-adjusted estimates and their precision (eg, 95% confidence interval). Make clear which confounders were adjusted for and why they were included | 11-13 |
| | | (*b*) Report category boundaries when continuous variables were categorized | - |

| | | | |
|---|---|---|---|
| | | (*c*) If relevant, consider translating estimates of relative risk into absolute risk for a meaningful time period | N/A |
| Other analyses | 17 | Report other analyses done—eg analyses of subgroups and interactions, and sensitivity analyses | N/A |
| **Discussion** | | | |
| Key results | 18 | Summarise key results with reference to study objectives | 25 |
| Limitations | 19 | Discuss limitations of the study, taking into account sources of potential bias or imprecision. Discuss both direction and magnitude of any potential bias | 25 |
| Interpretation | 20 | Give a cautious overall interpretation of results considering objectives, limitations, multiplicity of analyses, results from similar studies, and other relevant evidence | 25-26 |
| Generalisability | 21 | Discuss the generalisability (external validity) of the study results | 25-26 |
| **Other information** | | | |
| Funding | 22 | Give the source of funding and the role of the funders for the present study and, if applicable, for the original study on which the present article is based | 26 |

*Give information separately for cases and controls in case-control studies and, if applicable, for exposed and unexposed groups in cohort and cross-sectional studies.

### Supplementary Material 2. Consolidated criteria for reporting qualitative studies (COREQ): 32-item checklist with page numbers to indicate section of the article

| Checklist item | Questions to consider | Page number in article |
|---|---|---|
| **Domain 1: Research team and reflexivity** | | |
| ***Personal Characteristics*** | | |
| 1. Interviewer/facilitator | Which author/s conducted the interview or focus group? | 8 |
| 2. Credentials | What were the researcher's credentials? *E.g. PhD, MD* | 1 |
| 3. Occupation | What was their occupation at the time of the study? | 1 and 26 |
| 4. Gender | Was the researcher male or female? | 1 |
| 5. Experience and training | What experience or training did the researcher have? | 1 |
| ***Relationship with participants*** | | |
| 6. Relationship established | Was a relationship established prior to study commencement? | Not reported due to limited word count |
| 7. Participant knowledge of the interviewer | What did the participants know about the researcher? e.*g. personal goals, reasons for doing the research* | Not reported due to limited word count |
| 8. Interviewer characteristics | What characteristics were reported about the interviewer/facilitator? e.g. *Bias, assumptions, reasons and interests in the research topic* | Not reported due to limited word count |
| **Domain 2: study design** | | |
| ***Theoretical framework*** | | |
| 9. Methodological orientation and Theory | What methodological orientation was stated to underpin the study? *e.g. grounded theory, discourse analysis, ethnography, phenomenology, content analysis* | 9 |
| ***Participant selection*** | | |
| 10. Sampling | How were participants selected? *e.g. purposive, convenience, consecutive, snowball* | 8 and Table 1 |
| 11. Method of approach | How were participants approached? e.*g. face-to-face, telephone, mail, email* | 8 |
| 12. Sample size | How many participants were in the study? | 9 and Table 3 |
| 13. Non-participation | How many people refused to participate or dropped out? Reasons? | Table 1 |
| ***Setting*** | | |
| 14. Setting of data collection | Where was the data collected? e.*g. home, clinic, workplace* | 8 |
| 15. Presence of non-participants | Was anyone else present besides the participants and researchers? | N/A |
| 16. Description of sample | What are the important characteristics of the sample? *e.g. demographic data, date* | 9 and Table 3 |
| ***Data collection*** | | |
| 17. Interview guide | Were questions, prompts, guides provided by the authors? Was it pilot tested? | Supplementary Material 5 & 6 |
| 18. Repeat interviews | Were repeat interviews carried out? If yes, how many? | N/A |
| 19. Audio/visual recording | Did the research use audio or visual recording to collect the data? | 9 |
| 20. Field notes | Were field notes made during and/or after the interview or focus group? | N/A |
| 21. Duration | What was the duration of the interviews or focus group? | Table 1 |
| 22. Data saturation | Was data saturation discussed? | N/A |
| 23. Transcripts returned | Were transcripts returned to participants for comment and/or correction? | N/A |
| **Domain 3: analysis and findings** | | |
| ***Data analysis*** | | |

| | | |
|---|---|---|
| 24. Number of data coders | How many data coders coded the data? | 9 |
| 25. Description of the coding tree | Did authors provide a description of the coding tree? | Supplementary material 7 & 8 |
| 26. Derivation of themes | Were themes identified in advance or derived from the data? | 9 |
| 27. Software | What software, if applicable, was used to manage the data? | 9 |
| 28. Participant checking | Did participants provide feedback on the findings? | N/A |
| ***Reporting*** | | |
| 29. Quotations presented | Were participant quotations presented to illustrate the themes / findings? Was each quotation identified? e.*g. participant number* | Table 4 |
| 30. Data and findings consistent | Was there consistency between the data presented and the findings? | 13-24 |
| 31. Clarity of major themes | Were major themes clearly presented in the findings? | 13-24 |
| 32. Clarity of minor themes | Is there a description of diverse cases or discussion of minor themes? | 13-24 |

Reference: Tong A, Sainsbury P, Craig J. Consolidated criteria for reporting qualitative research (COREQ): a 32-item checklist for interviews and focus groups. Int J Qual Health Care. 2007;19(6):349-357.

### Supplementary Material 3. Assessment criteria for the delivery of essential components of standard best practice by quit status on the screening van and at four-week assessment[1]

| | Pre-quit | | Quit date | | <3 weeks post-quit | | >3 weeks post-quit | |
|---|---|---|---|---|---|---|---|---|
| **Assessment criteria** | **Screening Van** | **Four weeks** | **Screening Van** | **Four weeks** | **Screening Van** | **Four weeks** | **Screening Van** | **Four weeks** |
| **Assess current readiness and ability to quit** | ✓ | ✓ | | | | | | |
| **Confirm readiness and ability to quit** | | | ✓ | ✓ | | | | |
| **Confirm quit status and progress** | | | | | ✓ | ✓ | ✓ | ✓ |
| **Inform the patient about the treatment programme** | ✓ | | ✓ | | ✓ | | ✓ | |
| **Assess current smoking** | ✓ | ✓ | | | | | | |
| **Assess past quit attempts** | ✓ | ✓ | ✓ | ✓ | ✓ | ✓ | ✓ | ✓ |
| **Explain how tobacco dependence develops and assess level of nicotine dependence** | | ✓ | ✓ | ✓ | | | | |
| **Explain carbon monoxide (CO) monitoring and discuss results (previously taken as part of YLST) using the CO reading chart** | ✓ | ✓ | ✓ | ✓ | ✓ | ✓ | ✓ | ✓ |
| **Explain the importance of abrupt cessation and the 'not a puff' rule** | | ✓ | ✓ | ✓ | ✓ | ✓ | ✓ | ✓ |
| **Inform about withdrawal symptoms** | | ✓ | | | | | | |
| **Discuss withdrawal symptoms and cravings/urges to smoke and how to deal with them** | | | ✓ | ✓ | ✓ | ✓ | ✓ | ✓ |
| **Discuss stop smoking medications/NRT/e-cigarettes and provide appropriate 2 weeks supply** | ✓ | ✓ | ✓ | ✓ | ✓ | ✓ | ✓ | ✓ |
| **Set quit date** | | ✓ | | | | | | |
| **Advise on changing routine** | | | ✓ | ✓ | | | | |
| **Discuss any difficult situations experienced and methods of coping** | | | | | ✓ | ✓ | ✓ | ✓ |
| **Address any potential high-risk situations in the coming week** | | | ✓ | ✓ | ✓ | ✓ | ✓ | ✓ |
| **Prompt commitment from the patient** | | ✓ | ✓ | ✓ | ✓ | ✓ | ✓ | ✓ |
| **Discuss preparations and planning and provide summary** | ✓ | ✓ | ✓ | ✓ | ✓ | ✓ | ✓ | ✓ |
| **Positive communication and developing rapport** | ✓ | ✓ | ✓ | ✓ | ✓ | ✓ | ✓ | ✓ |
| **Theoretical maximum fidelity score for green (essential) primary assessment criteria** | 8 | 12 | 14 | 13 | 12 | 11 | 12 | 11 |

[1]Ticked boxes denote essential components of standard best practice that should be delivered at each time point.

### Supplementary Material 4. Assessment criteria for the delivery of essential components of the YESS Booklet intervention by booklet number at four-weeks[1]

| | | Booklet | | | | | |
|---|---|---|---|---|---|---|---|
| **Domain** | **Assessment criteria** | No heart damage; moderate lung damage | No heart damage; no lung damage | Heart damage; moderate lung damage | Heart damage; no lung damage | No heart damage; severe lung damage | Heart damage ; severe lung damage |
| ***Communication Brief for delivery Overall*** | Encouragement, motivation, understanding, and empathy | ✓ | ✓ | ✓ | ✓ | ✓ | ✓ |
| | Reflective listening | ✓ | ✓ | ✓ | ✓ | ✓ | ✓ |
| | Checking understanding/comprehension (e.g. talk back (a method of checking participants understanding by asking them stating it in their own words or explaining what something means)) | ✓ | ✓ | ✓ | ✓ | ✓ | ✓ |
| | Use lay language | ✓ | ✓ | ✓ | ✓ | ✓ | ✓ |
| | Presents YESS and YLST as professional and expert services. | ✓ | ✓ | ✓ | ✓ | ✓ | ✓ |
| | Explains role of SCP: that they are not a medical doctor and therefore the limits to the clinical advice that can/will be given by YESS smoking cessation practitioners and they cannot answer any clinical questions about results. | ✓ | ✓ | ✓ | ✓ | ✓ | ✓ |
| | Highlight that there is a phone number on the booklet if they have any queries that they wish to discuss with the YLST clinical team. | ✓ | ✓ | ✓ | ✓ | ✓ | ✓ |
| | Builds confidence in participant that they can quit | ✓ | ✓ | ✓ | ✓ | ✓ | ✓ |
| | Is sympathetic to, and acknowledges, the difficulties of quitting | ✓ | ✓ | ✓ | ✓ | ✓ | ✓ |
| | Relates results/risks and proposed benefits of quitting directly to the individual | ✓ | ✓ | ✓ | ✓ | ✓ | ✓ |
| | Emphasises immediate/short-term benefits of quitting regardless of scan result and importantly, for any age | ✓ | ✓ | ✓ | ✓ | ✓ | ✓ |
| ***Introduction to the booklet*** | Emphasise salience from outset (these are *your* images) | ✓ | ✓ | ✓ | ✓ | ✓ | ✓ |
| | Expectation setting | ✓ | ✓ | ✓ | ✓ | ✓ | ✓ |
| ***'Your lungs' page*** | Describe – describe lung images; describing light bits as bone, dark bits as tissue, and 'circled areas' as damaged areas | ✓ | ✓ | ✓ | ✓ | ✓ | ✓ |
| | Explain and clarify risk - clarify continued risks of smoking in context of CT result by using lay explanation of how smoking causes emphysema and how emphysema affects the participant's health, in relation to level of lung damage. | ✓ | ✓ | ✓ | ✓ | ✓ | ✓ |
| | Increase response efficacy - by explaining how stopping smoking has immediate benefits of limiting further damage (for those with damage) or preventing damage (for those without damage). | ✓ | ✓ | ✓ | ✓ | ✓ | ✓ |
| | Manage/reduce potential for over-reassurance as these participants have no lung damage | | ✓ | | ✓ | | |
| | Manage anxiety about the extent of smoking damage, including questions about this not being reversible | | | | | | |

| | | | | | | | |
|---|---|---|---|---|---|---|---|
| | Manage queries about a diagnosis/disclosure of emphysema (when they were unaware they had emphysema) and anxiety and concerns about it | | | | | | |
| ***'Your heart' page*** | Describe – describe heart images - orientating patients to other areas of the image that should not be confused with calcification of the heart (spine for example) | ✓ | ✓ | ✓ | ✓ | ✓ | ✓ |
| | Explain and clarify risk - clarify continued risks of smoking in context of CT result by explaining that smoking is one likely cause of their artery narrowing and hardening. | ✓ | ✓ | ✓ | ✓ | ✓ | ✓ |
| | Set this in the context of other causes so as not to over-exaggerate role of smoking, which participants may discredit. | ✓ | ✓ | ✓ | ✓ | ✓ | ✓ |
| | Increase response efficacy - by explaining how stopping smoking has immediate benefits of limiting further damage (for those with damage) or preventing damage (for those without damage). | ✓ | ✓ | ✓ | ✓ | ✓ | ✓ |
| | Manage/reduce potential for over-reassurance as these participants have 'no' heart damage | ✓ | ✓ | | | ✓ | |
| | Manage queries about other non-smoking causes of heart damage | | | | | | ✓ |
| | Manage scepticism about the role of smoking in future heart disease | | | | | | |
| | Manage anxiety and concerns about disclosure of coronary artery calcification | | | | | | |
| ***'How stopping smoking will help you' page*** | Increase response efficacy by emphasising the short-term benefits of stopping smoking and relating this directly to the individual | ✓ | ✓ | ✓ | ✓ | ✓ | ✓ |
| | Help patient envision how their health will improve over the first year (focusing less on the longer term). | ✓ | ✓ | ✓ | ✓ | ✓ | ✓ |
| | Use lay explanations to show how these health benefits accumulate and work to achieve significant reductions in risk over the following years. | ✓ | ✓ | ✓ | ✓ | ✓ | ✓ |
| | Emphasise relevance for older age | ✓ | ✓ | ✓ | ✓ | ✓ | ✓ |
| ***Closing Session*** | Provide a positive close and reiterate key messages set out at the beginning | ✓ | ✓ | ✓ | ✓ | ✓ | ✓ |
| | Check patients understanding and comprehension of leaflet. | ✓ | ✓ | ✓ | ✓ | ✓ | ✓ |
| | Further safety net against potential over reassurance for those with clear results | | | | | | |
| Theoretical maximum fidelity score for essential assessment criteria | | 27 | 28 | 26 | 27 | 27 | 27 |

[11]Ticked boxes denote essential components that should be delivered by booklet type

### Supplementary Material 5. Topic guides for interviews with SCPs

SCP 4 week interview topic guide

***Rationale:*** *to gain views on (a) intervention usage (b) aspects perceived to influence change (c) contextual barriers and facilitators (d) embedding the intervention into practice (e) potential contamination (f) other*

#### Background

- Welcome and thanks; introduce yourself
- Give a brief background on the purpose of the interview
- Explain what will happen in the interview
- Reaffirm the interviewees right to withdraw and ensure that they are comfortable
- Reaffirm that the interviewee is happy for the interview to audio-recorded and anonymised quotes could be used in future reports or publications Check that the interviewee understands the reason for the interview and the role of the researcher. Provide an opportunity for the interviewee to ask any questions

#### A. Intervention usage

- **Almost a bit like a story, can you talk me through the process of the study please?**
  - Do you find this varies a lot dependent on the participant?
    - If so, why do you think that might be?

#### B. Aspects perceived to influence change

- **There are many aspects of the intervention, what do you feel the main ones are?**
  - Do you think there is one aspect that is more important?
    - Why do you think it's that?
  - Do you think there are any aspects that are less important?
    - Why do you think that?
- **There are many key messages we aim to get across in the intervention, what do you feel are the main ones?**
  - Do you think there are any that is more important?
  - Why do you think it's that?
  - Do you think there are any that are almost left behind?
- **Thinking about participants now – which aspects do you think they respond to more?**
  - Why? How?
  - Are there any parts of the intervention that tend to raise a few questions or talking points? (Why? How?)
- **Thinking about participants again – which messages do you think they respond to more?**
  - Why? How?
  - Are there any messages that tend of raise a few questions or talking points?
- **Have you had any participants that have been shocked by their results? How have you dealt with this?**
- **Do you feel like participants understood what you were trying to get across?**

#### C. Contextual barriers and facilitators

- **How did you feel talking to participants about smoking cessation and their heart and lung health?**
  - Did you find that participants were amenable to the information you were giving them?

SCP 12 week topic guide

***Rationale:** to gain views on (a) completing follow-ups (b) contextual barriers and facilitators (c) potential contamination (d) other*

### Background

- Welcome and thanks; introduce yourself
- Give a brief background on the purpose of the interview
- Explain what will happen in the interview
- Reaffirm the interviewees right to withdraw and ensure that they are comfortable
- Reaffirm that the interviewee is happy for the interview to audio-recorded and anonymised quotes could be used in future reports or publications
- Check that the interviewee understands the reason for the interview and the role of the researcher
- Provide an opportunity for the interviewee to ask any questions

#### A. Completing follow-ups

- **Can you tell me a little bit about how the follow-ups run?**
    - Do you find this varies a lot dependent on the participant?
        - If so, why do you think that might be?
- **How do participants react to completing the questionnaire?**
    - Does this vary my participant/age/allocation?
- **Do you think completing the questionnaire has an effect on the participants (i.e. the questionnaire influencing their quit motivation)**
- **Do any of the questions raise questions or act as a point of discussion?**

#### B. Contextual barriers and facilitators

- **How did you feel talking to participants about smoking cessation and their heart and lung health?**
    - Did you find that participants were amenable to the information you were giving them?
- **During the follow-ups did participants discuss any barriers for smoking cessation to you?**
    - What were they?
        - What explanations did they provide for these?
    - What do you think is the most important barrier that we could try and break?
- **During the follow-ups did participant discuss any facilitators for smoking cessation to you?**
    - What were they?
        - What explanations did they provide for these?
    - What do you think is the most important facilitator that we could try and include?

#### C. Potential contamination

- **Did participants talk to you about receiving smoking cessation or lung health information from anywhere else?**
    - What type of information was this?
    - Where did it come from?
    - Do you think this information had had any impact on their opinions of smoking cessation before they came to talk to you?
- **Did participants mention their awareness of any stop smoking or lung health awareness campaigns?**
    - What type of campaign was this?
    - Where did it come from?

  - Do you think this information had had any impact on their opinions of smoking cessation before they came to talk to you?
- **Have any participants mention that they knew anyone else who was involved in the study?**
  - What did they mention about this?
  - Do you think this had an impact on their smoking cessation journey or opinions?

#### D. Other

- **Do you have any other questions or points that you would like to make?**

### SCP 52 week Interview Topic Guide

***Rationale:*** *to gain views on (a) completing follow-ups (b) contextual barriers and facilitators (c) potential contamination (d) other*

#### Background

- Welcome and thanks; introduce yourself
- Give a brief background on the purpose of the interview
- Explain what will happen in the interview
- Reaffirm the interviewees right to withdraw and ensure that they are comfortable
- Reaffirm that the interviewee is happy for the interview to audio-recorded and anonymised quotes could be used in future reports or publications
- Check that the interviewee understands the reason for the interview and the role of the researcher
- Provide an opportunity for the interviewee to ask any questions

#### A. Completing follow-ups

- **Can you tell me a little bit about how the follow-ups run?**
  - Do you find this varies a lot dependent on the participant?
    - If so, why do you think that might be?
- **How do participants react to completing the questionnaire?**
  - Does this vary my participant/age/allocation?
- **Do you think completing the questionnaire has an effect on the participants (i.e. the questionnaire influencing their quit motivation)**
- **Do any of the questions raise questions or act as a point of discussion?**

#### B. Contextual barriers and facilitators

- **How did you feel talking to participants about smoking cessation and their heart and lung health?**
  - Did you find that participants were amenable to the information you were giving them?
- **During the follow-ups did participants discuss any barriers for smoking cessation to you?**
  - What were they?
    - What explanations did they provide for these?
  - What do you think is the most important barrier that we could try and break?
- **During the follow-ups did participant discuss any facilitators for smoking cessation to you?**
  - What were they?
    - What explanations did they provide for these?
  - What do you think is the most important facilitator that we could try and include?

#### C. Potential contamination

- **Did participants talk to you about receiving smoking cessation or lung health information from anywhere else?**
  - What type of information was this?
  - Where did it come from?
  - Do you think this information had had any impact on their opinions of smoking cessation before they came to talk to you?
- **Did participants mention their awareness of any stop smoking or lung health awareness campaigns?**
  - What type of campaign was this?
  - Where did it come from?
  - Do you think this information had had any impact on their opinions of smoking cessation before they came to talk to you?
- **Have any participants mention that they knew anyone else who was involved in the study?**
  - What did they mention about this?
  - Do you think this had an impact on their smoking cessation journey or opinions?

#### D. Other

- **Do you have any other questions or points that you would like to make?**

### Supplementary Material 6. Topic guides for interviews with trial participants

CONTROL participants 4-week Interview Topic Guide

***Rationale:*** *to gain views on (a) potential study contamination (b) influence of COVID-19 (b) other*

#### Background

- Welcome and thanks; introduce yourself
- Give a brief background on the purpose of the interview
- Explain what will happen in the interview
- Reaffirm the interviewees right to withdraw and ensure that they are comfortable
- Reaffirm that the interviewee is happy for the interview to audio-recorded and anonymised quotes could be used in future reports or publications
- Check that the interviewee understands the reason for the interview and the role of the researcher
- Provide an opportunity for the interviewee to ask any questions

#### A. Potential Study Contamination

- **Since being involved in the study have you received any further smoking cessation or lung health information?**
    - If so, what and how?
        - Do you think this additional information has had any impact on how you feel about smoking and your lung health?
- **Since being involved in the study have you received any further smoking cessation or lung health support?**
    - If so, what and how? Do you think this additional support has had any impact on how you feel about smoking and your lung health?
- **Since being involved in the study have you been aware of any stop smoking campaigns? (these could be in the media on TV and radio, or locally)**
    - If so, which campaigns were they?
        - Can you remember any of the messages from the campaign? Or any particular topics covered?
    - What impact if any did the campaign have on you?
- **Since being part of the study are you aware of anyone else who took part in the study?**
    - If so, did you discuss the study with them?
    - Are you aware if they were shown pictures of their scans too?
        - Did you talk about their results with them?
        - How did it feel talking to them about it?
- **Did you talk to anybody else about being part of the study?**
    - Why do you feel that you did this?
    - What did you discuss with them?

**B. Influence of COVID-19** *(If participant brings COVID or the impact of the pandemic up earlier in this interview then discuss these points then as appropriate, if not, then leave to the end)*

- How do you feel about the COVID-19 pandemic?
- What, if any, impact has it had on how you feel about smoking/continuing to smoke/trying to stop?
- Have your smoking habits changed at all since the pandemic started in March 2020?
- How do you feel it may have affected your journey to stopping smoking/cutting down?

#### C. Other

- **Do you have any other questions or points that you would like to make?**

INTERVENTION participants 4-week Interview Topic Guide

***Rationale:*** *to gain views on (a) intervention usage (b) intervention comprehension (c) influence of intervention on knowledge, beliefs and behaviours (d) intervention acceptability (e) potential study contamination (f) influence of COVID-19 (g) other*

### Background

- Welcome and thanks; introduce yourself
- Give a brief background on the purpose of the interview
- Explain what will happen in the interview
- Reaffirm the interviewees right to withdraw and ensure that they are comfortable
- Reaffirm that the interviewee is happy for the interview to audio-recorded and anonymised quotes could be used in future reports or publications
- Check that the interviewee understands the reason for the interview and the role of the researcher
- Provide an opportunity for the interviewee to ask any questions

#### A. Intervention Usage

- **If it's ok with you, I'd like to start with you telling me what happened? Should we start with when you were invited to join the study?** *(Tailor follow-up questions to telephone-based intervention delivery where appropriate)*
    - What were the processes that you went through?
    - What did you discuss with the advisor?
    - Did the advisor have any materials?
        - Did you talk about these materials?
    - Were you given any materials to take home?/Did you receive your booklet when it was sent to you?
- **Why did you decide to go and see the smoking cessation practitioner at your lung health check?**

#### B. Intervention Comprehension

- **And if it's ok now, can you tell me a little bit about what it all meant? What were the take home messages for you from the intervention?**
    - What did your results mean to you?
    - What do you think the advisor was trying to tell you?
    - What do you think about quitting smoking?

#### C. Influence of intervention on knowledge, beliefs and behaviours

- **As you have just described earlier there were a few different parts to the intervention, do you think that any were more important than others?** *(Tailor follow-up questions to telephone-based intervention delivery where appropriate)*
    - Do you think any had more of an influence on how you feel about smoking or stopping smoking?
    - Do you think any had more of an influence on how you feel about your health?
    - Has completing the intervention with the advisor made you think or feel differently in any way? This could be about your health or smoking at all.
        - What parts of the intervention made you feel like this?
        - Why you think this part of the intervention made you feel like this?
- **Since receiving the intervention have you made any changes to stop smoking?**
    - Why did you make these changes?
    - Do you feel like you will be able to keep up these changes?
        - Do you think anything else could be done to help you with this? If so, what and how?
    - If not, do you mind me asking why you have not made any changes?
- **Since receiving the intervention have you made any changes to improve your health?**
    - Why did you make these changes?

- Do you feel like you will be able to keep up these changes?
  - Do you think anything else could be done help you with this? If so, what and how?
- If not, do you mind me asking why you have not made any changes?

#### D. Intervention Acceptability

- **How did you feel about completing the intervention?** *(Tailor follow-up questions to telephone-based intervention delivery where appropriate)*
  - How did you feel about talking to the advisor?
  - How easy was it for you to trust the advisor?
  - How did you feel receiving your results?
  - How did you feel about the information that was given to you by the advisor?
  - Did any of the information given shock or surprise you? If so, what? And why do you think it shocked you?
- **The intervention was given to you approximately 4 weeks after your lung screening appointment in a community van (or home), how did you feel about receiving it then?**
  - How did you feel about it being in the van?
  - How did you feel taking part in the lung health check?
    - Do you think that made a difference about how you feel about smoking and the stop smoking advice you have received?
  - How did you feel about being approached to talk about stopping smoking when you went for your lung health check?

#### E. Potential Study Contamination

- **Since being involved in the study have you received any further smoking cessation or lung health information?**
  - If so, what and how?
    - Do you think this additional information has had any impact on how you feel about smoking and your lung health?
- **Since being involved in the study have you received any further smoking cessation or lung health support?**
  - If so, what and how?
    - Do you think this additional support has had any impact on how you feel about smoking and your lung health?
- **Since being involved in the study have you been aware of any stop smoking campaigns? (these could be in the media on TV and radio, or locally)**
  - If so, which campaigns were they?
    - Can you remember any of the messages from the campaign? Or any particular topics covered?
  - What impact if any did the campaign have on you?
- **Since being part of the study are you aware of anyone else who took part in the study?**
  - If so, did you discuss the study with them?
  - Are you aware if they were shown pictures of their scans too?
    - Did you talk about their results with them?
    - How did it feel talking to them about it?
- **Did you talk to anybody else about being part of the study?**
  - Why do you feel that you did this?
  - What did you discuss with them?

#### F. Influence of COVID-19 *(If participant brings COVID or the impact of the pandemic up earlier in this interview then discuss these points then as appropriate, if not, then leave to the end)*

- How do you feel about the COVID-19 pandemic?
- What, if any, impact has it had on how you feel about smoking/continuing to smoke/trying to stop?
- Have your smoking habits changed at all since the pandemic started in March?
- How do you feel it may have affected your journey to stopping smoking/cutting down?

#### G. Other

- **Do you have any other questions or points that you would like to make?**

CONTROL participants 12-week Interview Topic Guide

***Rationale:*** *to gain views on (a) potential study contamination (b) influence of COVID-19 (c) other*

### Background

- Welcome and thanks; introduce yourself
- Give a brief background on the purpose of the interview
- Explain what will happen in the interview
- Reaffirm the interviewees right to withdraw and ensure that they are comfortable
- Reaffirm that the interviewee is happy for the interview to audio-recorded and anonymised quotes could be used in future reports or publications
- Check that the interviewee understands the reason for the interview and the role of the researcher
- Provide an opportunity for the interviewee to ask any questions

### A. Potential Study Contamination

- **Since being involved in the study have you received any further smoking cessation or lung health information?**
    - If so, what and how?
        - Do you think this additional information has had any impact on how you feel about smoking and your lung health?
- **Since being involved in the study have you received any further smoking cessation or lung health support?**
    - If so, what and how?
        - Do you think this additional support has had any impact on how you feel about smoking and your lung health?
- Since being involved in the study have you been aware of any stop smoking campaigns? (these could be in the media on TV and radio, or locally)
    - If so, which campaigns were they?
        - Can you remember any of the messages from the campaign? Or any particular topics covered?
    - What impact if any did the campaign have on you?
- **Since being part of the study are you aware of anyone else who took part in the study?**
    - If so, did you discuss the study with them?
    - Are you aware if they were shown pictures of their scans too?
        - Did you talk about their results with them?
        - How did it feel talking to them about it?
- **Did you talk to anybody else about being part of the study?**
    - Why do you feel that you did this?
    - What did you discuss with them?

**B. Influence of COVID-19** *(If participant brings COVID or the impact of the pandemic up earlier in this interview then discuss these points then as appropriate, if not, then leave to the end)*

- How do you feel about the COVID-19 pandemic?
- What, if any, impact has it had on how you feel about smoking/continuing to smoke/trying to stop?
- Have your smoking habits changed at all since the pandemic started in March 2020?
- How do you feel it may have affected your journey to stopping smoking/cutting down?

### C. Other

- **Do you have any other questions or points that you would like to make?**

INTERVENTION participants 12-week Interview Topic Guide

***Rationale:** to gain views on (a) intervention usage (b) intervention comprehension (c) influence of intervention on knowledge, beliefs and behaviours (d) intervention acceptability (e) potential study contamination (f) influence of COVID-19 (g) other*

### Background

- Welcome and thanks; introduce yourself
- Give a brief background on the purpose of the interview
- Explain what will happen in the interview
- Reaffirm the interviewees right to withdraw and ensure that they are comfortable
- Reaffirm that the interviewee is happy for the interview to audio-recorded and anonymised quotes could be used in future reports or publications
- Check that the interviewee understands the reason for the interview and the role of the researcher
- Provide an opportunity for the interviewee to ask any questions

### A. Intervention Usage

- **I know it's quite a few weeks ago now but if it's ok with you, I'd like to start with you telling me what happened? Should we start with when you were invited to join the study?** *(Tailor follow-up questions to telephone-based intervention delivery where appropriate)*
    - What were the processes that you went through?
    - What did you discuss with the advisor?
    - Did the advisor have any materials?
        - Did you talk about these materials?
    - Were you given any materials to take home?/Did you receive your booklet when it was sent to you?
- **Why did you decide to go and see the smoking cessation practitioner at your lung health check?**

### B. Intervention Comprehension

- **And if it's ok now, can you tell me a little bit about what it all meant?**
    - What did your results mean to you?
    - What do you think the advisor was trying to tell you?
    - What do you think about quitting smoking?

### C. Influence of intervention on knowledge, beliefs and behaviours

- **As you have just described earlier there were a few different parts to the intervention, do you think that any were more important than others?** *(Tailor follow-up questions to telephone-based intervention delivery where appropriate)*
    - Do you think any had more of an influence on how you feel about smoking or stopping smoking?
    - Do you think any had more of an influence on how you feel about your health?
    - Has completing the intervention with the advisor made you think or feel differently in any way? This could be about your health or smoking at all.
        - What parts of the intervention made you feel like this?
        - Why you think this part of the intervention made you feel like this?
- **Since receiving the intervention have you made any changes to stop smoking?**
    - Why did you make these changes?
    - Do you feel like you will be able to keep up these changes?
        - Do you think anything else could be done to help you with this? If so, what and how?
    - If not, do you mind me asking why you have not made any changes?

- **Since receiving the intervention have you made any changes to improve your health?**
  - Why did you make these changes?
  - Do you feel like you will be able to keep up these changes?
  - Do you think anything else could be done help you with this? If so, what and how?
  - If not, do you mind me asking why you have not made any changes?
- **Recently you completed a follow-up questionnaire over the phone/at home with a colleague of mine, did any of the questions in particular make you stop and think?**
  - Which questions were they? Why do you think they did this?

**D. Intervention Acceptability**

- **How did you feel about completing the intervention?** *(Tailor follow-up questions to telephone-based intervention delivery where appropriate)*
  - How did you feel about talking to the advisor?
  - How easy was it for you to trust the advisor?
  - How did you feel receiving your results?
  - How did you feel about the information that was given to you by the advisor?
  - Did any of the information given shock or surprise you? If so, what? And why do you think it shocked you?
- **The intervention was given to you approximately 4 weeks after your lung screening appointment in a community van (or home, or over the phone), how did you feel about receiving it then?**
  - How did you feel about it being in the van?
  - How did you feel taking part in the lung health check?
    - Do you think that made a difference about how you felt about smoking and the stop smoking advice you have received?
  - How did you feel about being approached to talk about stopping smoking when you went for your lung health check?
- **As part of the study you recently completed a follow-up over the telephone/at home, how did you feel about doing this?**

**E. Potential Study Contamination**

- **Since being involved in the study have you received any further smoking cessation or lung health information?**
  - If so, what and how?
    - Do you think this additional information has had any impact on how you feel about smoking and your lung health?
- **Since being involved in the study have you received any further smoking cessation or lung health support?**
  - If so, what and how?
    - Do you think this additional support has had any impact on how you feel about smoking and your lung health?
- **Since being involved in the study have you been aware of any stop smoking campaigns? (these could be in the media on TV and radio, or locally)**
  - If so, which campaigns were they?
    - Can you remember any of the messages from the campaign? Or any particular topics covered?
  - What impact if any did the campaign have on you?
- **Since being part of the study are you aware of anyone else who took part in the study?**
  - If so, did you discuss the study with them?
  - Are you aware if they were shown pictures of their scans too?
    - Did you talk about their results with them?
    - How did it feel talking to them about it?
- **Did you talk to anybody else about being part of the study?**
  - Why do you feel that you did this?
  - What did you discuss with them?

**F. Influence of COVID-19** *(If participant brings COVID or the impact of the pandemic up earlier in this interview then discuss these points then as appropriate, if not, then leave to the end)*

- **How do you feel about the COVID-19 pandemic?**
- What, if any, impact has it had on how you feel about smoking/continuing to smoke/trying to stop?
- Have your smoking habits changed at all since the pandemic started in March?
- How do you feel it may have affected your journey to stopping smoking/cutting down?

**G. Other**

- **Do you have any other questions or points that you would like to make?**

<u>CONTROL participants 52-week interview topic guide</u>

***Rationale:*** *to gain views on (a) potential study contamination (b) influence of COVID-19 (c) other*

**Background**

- Welcome and thanks; introduce yourself
- Give a brief background on the purpose of the interview
- Explain what will happen in the interview
- Reaffirm the interviewees right to withdraw and ensure that they are comfortable
- Reaffirm that the interviewee is happy for the interview to audio-recorded and anonymised quotes could be used in future reports or publications
- Check that the interviewee understands the reason for the interview and the role of the researcher
- Provide an opportunity for the interviewee to ask any questions

**A. Potential Study Contamination**

- **Since being involved in the study have you received any further smoking cessation or lung health information?**
  - If so, what and how?
    - Do you think this additional information has had any impact on how you feel about smoking and your lung health?
- **Since being involved in the study have you received any further smoking cessation or lung health support?**
  - If so, what and how?
    - Do you think this additional support has had any impact on how you feel about smoking and your lung health?
- Since being involved in the study have you been aware of any stop smoking campaigns? (these could be in the media on TV and radio, or locally)
  - If so, which campaigns were they?
    - Can you remember any of the messages from the campaign? Or any particular topics covered?
  - What impact if any did the campaign have on you?
- **Since being part of the study are you aware of anyone else who took part in the study?**
  - If so, did you discuss the study with them?
  - Are you aware if they were shown pictures of their scans too?
    - Did you talk about their results with them?
    - How did it feel talking to them about it?
- **Did you talk to anybody else about being part of the study?**
  - Why do you feel that you did this?
  - What did you discuss with them?

**B. Influence of COVID-19** *(If participant brings COVID or the impact of the pandemic up earlier in this interview then discuss these points then as appropriate, if not, then leave to the end)*

- How do you feel about the COVID-19 pandemic?

- What, if any, impact has it had on how you feel about smoking/continuing to smoke/trying to stop?
- Have your smoking habits changed at all since the pandemic started in March 2020?
- How do you feel it may have affected your journey to stopping smoking/cutting down?

**C. Other**

- **Do you have any other questions or points that you would like to make?**

INTERVENTION participants 52 WEEK Interview Topic Guide

***Rationale:*** *to gain views on (a) intervention usage (b) intervention comprehension (c) influence of intervention on knowledge, beliefs and behaviours (d) intervention acceptability (e) potential study contamination (f) influence of COVID (g) other*

**Background**

- Welcome and thanks; introduce yourself
- Give a brief background on the purpose of the interview
- Explain what will happen in the interview
- Reaffirm the interviewees right to withdraw and ensure that they are comfortable
- Reaffirm that the interviewee is happy for the interview to audio-recorded and anonymised quotes could be used in future reports or publications
- Check that the interviewee understands the reason for the interview and the role of the researcher
- Provide an opportunity for the interviewee to ask any questions

**A. Intervention Usage**

- **I know it's approximately a year ago now but can we start by you talking me thorugh what you can remember happening please?? Should we start with when you were invited to join the study?** *(Tailor follow-up questions to telephone-based intervention delivery where appropriate)*
    - What were the processes that you went through?
    - What did you discuss with the advisor?
    - Did the advisor have any materials?
        - Did you talk about these materials?
    - Were you given any materials to take home?/Did you receive your booklet when it was sent to you?
- **Why did you decide to go and see the smoking cessation practitioner at your lung health check?**

**B. Intervention Comprehension**

- **And if it's ok now, can you tell me a little bit about what it all meant?**
    - What did your results mean to you?
    - What do you think the advisor was trying to tell you?
    - What do you think about quitting smoking?

**C. Influence of intervention on knowledge, beliefs and behaviours**

- **As you have just described earlier there were a few different parts to the intervention, do you think that any were more important than others?** *(Tailor follow-up questions to telephone-based intervention delivery where appropriate)*
    - Do you think any had more of an influence on how you feel about smoking or stopping smoking?
    - Do you think any had more of an influence on how you feel about your health?

- Has completing the intervention with the advisor made you think or feel differently in any way? This could be about your health or smoking at all.
  - What parts of the intervention made you feel like this?
  - Why you think this part of the intervention made you feel like this?

- **Since receiving the intervention have you made any changes to stop smoking?**
  - Why did you make these changes?
  - Do you feel like you will be able to keep up these changes?
    - Do you think anything else could be done to help you with this? If so, what and how?
  - If not, do you mind me asking why you have not made any changes?
- **Since receiving the intervention have you made any changes to improve your health?**
  - Why did you make these changes?
  - Do you feel like you will be able to keep up these changes?
    - Do you think anything else could be done help you with this? If so, what and how?
  - If not, do you mind me asking why you have not made any changes?
- **Recently you completed a follow-up questionnaire over the phone/at home with a colleague of mine, did any of the questions in particular make you stop and think?**
  - Which questions were they? Why do you think they did this?

**D. Intervention Acceptability**

- **How did you feel about completing the intervention?** *(Tailor follow-up questions to telephone-based intervention delivery where appropriate)*
  - How did you feel about talking to the advisor?
  - How easy was it for you to trust the advisor?
  - How did you feel receiving your results?
  - How did you feel about the information that was given to you by the advisor?
  - Did any of the information given shock or surprise you? If so, what? And why do you think it shocked you?
- **The intervention was given to you approximately 4 weeks after your lung screening appointment in a community van (or home), how did you feel about receiving it then?**
  - How did you feel about it being in the van?
  - How did you feel taking part in the lung health check?
    - Do you think that made a difference about how you feelt about smoking and the stop smoking advice you received?
  - How did you feel about being approached to talk about stopping smoking when you went for your lung health check?
- **As part of the study you recently completed a follow-up over the telephone/at home, how did you feel about doing this?**

**E. Potential Study Contamination**

- **Since being involved in the study have you received any further smoking cessation or lung health information?**
  - If so, what and how?
    - Do you think this additional information has had any impact on how you feel about smoking and your lung health?
- **Since being involved in the study have you received any further smoking cessation or lung health support?**
  - If so, what and how?
    - Do you think this additional support has had any impact on how you feel about smoking and your lung health?
- **Since being involved in the study have you been aware of any stop smoking campaigns? (these could be in the media on TV and radio, or locally)**
  - If so, which campaigns were they?
    - Can you remember any of the messages from the campaign? Or any particular topics covered?
  - What impact if any did the campaign have on you?
- **Since being part of the study are you aware of anyone else who took part in the study?**
  - If so, did you discuss the study with them?

    - Are you aware if they were shown pictures of their scans too?
        - Did you talk about their results with them?
        - How did it feel talking to them about it?
- **Did you talk to anybody else about being part of the study?**
    - Why do you feel that you did this?
    - What did you discuss with them?

**F. Influence of COVID-19** *(If participant brings COVID or the impact of the pandemic up earlier in this interview then discuss these points then as appropriate, if not, then leave to the end)*

- How do you feel about the COVID-19 pandemic?
- What, if any, impact has it had on how you feel about smoking/continuing to smoke/trying to stop?
- Have your smoking habits changed at all since the pandemic started in March 2020?
- How do you feel it may have affected your journey to stopping smoking/cutting down?

**G. Other**

- **Do you have any other questions or points that you would like to make?**

### Supplementary material 7: SCP interview coding framework (combined across timepoints)

| Major Theme | Sub Theme | Definition | Code No. |
|---|---|---|---|
| **SCP's background**<br><br>*Consider context* | Previous experiences with other smoking cessation services | Any discussions or points in reference to their previous experiences practicing in other smoking cessation services and their potential impact on the intervention/study. This can include both positive/negative experiences. | **1a** |
| | Other SCP's characteristics | Any discussions or points in reference to any other relevant contextual information about the SCPs. For example, information about SCP's smoking behaviours or professional background other than practicing as a smoking cessation advisor. | **1b** |
| **Training**<br><br>*Consider context, theory testing* | Trial-related training for enhanced intervention delivery | Any discussions or points in reference to the training provided by the YESS or YLST study team for enhanced (booklet & script) intervention delivery. All aspects of the **training**, including acceptability of training should be coded here. Acceptability of delivering the intervention with participants from the perspective of the SCP should be coded at 5a. | **2a** |
| | Training for standard best practice smoking cessation support – NCSCT training | Any discussions or points in reference to the training provided to help them deliver the smoking cessation support. This also include NCSCT training provided pre-trial or for the trial. All aspects of the **training**, including acceptability of training should be coded here. Acceptability of delivering the standard best practice with participants from the perspective of the SCP should be coded at 4b. | **2b** |
| | Trial-related training for other trial-specific components | Any discussions or points in reference to the training provided by the study to help them deliver other trial-specific components (e.g., taking consent, filling out the trial questionnaires, explaining randomisation allocation, etc.). Acceptability of delivering the trial specific components should be coded at 9a. | **2c** |
| | Support for SCPs provided by the trial study team | Any discussions or points in reference to any other types of support provided by the study team to help with their practice (both before and during the study process). Code here when the support was not provided as part of trial-related training. | **2d** |
| **Enhanced intervention comprehension**<br><br>*Consider intervention fidelity, exposure, and context* | | Any discussion or points in reference to **SCP's perception of participants' understanding** of the intervention and their personal results. Also code here if the SCPs refer to examples of when the participants described understanding (or not) the enhanced intervention. Important to consider:<br>• barriers/enablers to intervention comprehension (e.g. literacy issues)<br>• any questions raised by participants that SCP referred to during the interview | **3** |
| **Intervention acceptability**<br><br>*Consider intervention fidelity, exposure, context, and recruitment and retention* | Acceptability of enhanced intervention delivery | Any discussion or points in reference to the acceptability of delivering the booklet intervention, i.e., how SCPs felt delivering the booklet and giving participants their personal results. Important to consider any barriers/facilitators to booklet intervention delivery. Impact of COVID-19 on intervention acceptability should be coded at 9c. | **4a** |
| | Acceptability of standard best practice smoking cessation support delivery | Any discussion or points in reference to the acceptability of delivering the co-located standard best practice smoking cessation support, i.e., how they felt delivering smoking cessation support on the screening van and across the 12-week program. Important to consider any barriers/facilitators to smoking cessation support delivery. Impact of COVID-19 on intervention acceptability should be coded at 9c. | **4b** |
| | Practicality for implementation | Any discussion or points in reference to the practicality of delivering the co-located SBP smoking cessation service and | **4c** |

| | | | |
|---|---|---|---|
| | | enhanced booklet intervention for future implementation. Consider feasibility of delivering SC support within lung cancer screening, having a co-located services and feasibility of the booklet in the future. Also code here when SCPs describe potential barriers/facilitators to feasibly delivering these components. | |
| | Perception of participants' attitudes towards provided service | Any discussion or points in relation to **SCP's perception of participant's acceptability** of both standard best practice and the enhanced intervention. SCPs may draw on examples of participant reactions to intervention components or discussions they had with participants in relation to how they felt about different elements of the YESS. Including but not exclusive to:<br>• participants' reactions to being offered smoking cessation support at their lung health check appointment<br>• participants' views on enhanced intervention components<br>• participants' views on their conversation with the SCP/views on SCP role/views on ongoing regular contact with SCP<br>• participants' views on acceptability of co-located services<br>• participants' views regarding support coming to an end<br>Try to separate attitudes towards the pre-YESS (i.e., LHC and co-located support) from the YESS (i.e., consented at 4 weeks to 12 weeks). Proxy views about intervention comprehension should be coded at 3 | **4d** |
| **Influence of enhanced intervention**<br><br>*Consider theory testing, intervention fidelity, exposure, and context* | Influence of personalised risk information from enhanced intervention | Any discussion or points in reference to the impact or influence that the personalised booklet and scripted behavioural support from SCPs has had on participant's current smoking cessation/quit attempt as provided as part of the trial. Code here when they discussed how they delivered the booklet intervention and handled different situations during booklet intervention delivery, for example, how they handled participant's emotive reactions towards the booklet. Also code here when SCPs discussed participants' threat response (either high, low, or no threat) to risk information provided as part of the trial. | **5a** |
| | Self-efficacy | Any discussion or points that demonstrate previous, current or changes in participants self-efficacy beliefs in relation to quitting smoking **as influenced by enhanced intervention (booklet + script)**, defined as an individual's belief in their ability/capability to carry out a behaviour/goal successfully. Code here if the SCP describes how they went about building self-efficacy beliefs and how participants responded/showed evidence of self-efficacy beliefs being built. Reflections on the training to build self-efficacy beliefs should be coded at 1b. | **5b** |
| | Response-efficacy | Any discussion or points that demonstrate previous, current or changes in participants response-efficacy beliefs in relation to quitting smoking as influenced by enhanced intervention (booklet + script), defined as an individual's belief in whether the recommended behaviour (stopping smoking) will address the threat (i.e. will stopping smoking improve their health or reduce health risks for them). | **5c** |
| **Influence of standard best practice smoking cessation support** | Influence of NRT products | Any discussion or points in reference to the impact or influence that NRT products have had on participants' **current** smoking cessation/quit attempt as provided as part of the trial. Also code here when SCP discussed how participants used the NRT products; for example, when SCP discussed how participants | **6a** |

| | | | |
|---|---|---|---|
| *Consider theory testing, intervention fidelity, exposure, and context* | | were moved down in term of NRT dosages. Try to tease out influence of NRT products on participants' SE and RE beliefs. | |
| | Influence of CO validation monitoring | Any discussion or points in reference to the impact or influence that conducting CO validation monitoring had on participant's current smoking cessation/quit attempt as part of the trial. Try to tease out influence of CO validation on participants' SE and RE beliefs. | **6b** |
| | Contact with SCPs | Any discussion or points in reference to how frequency of contact with personalised support from SCPs influenced participants' smoking cessation. Code here when they discussed how they delivered SBP and handled different situations during smoking cessation support delivery, for example, how they handled participant's resistance towards smoking cessation support on SV. Try to tease out influence of SCP contact on participants' SE and RE beliefs. | **6c** |
| **Contextual facilitators or barriers for participants to quit smoking**<br><br>*Consider context, and recruitment and retention* | Influence of lung health check | Any discussion or points in reference to the impact or influence that the lung health check has had on participant's smoking behaviour. Consider lung health checks as a 'teachable moment'. | **7a** |
| | Social influences on smoking and smoking cessation | Any discussion or points in reference to the social influences on participant's smoking behaviour from their social environment (i.e. when the SCPs describe participant social networks as influences on quitting smoking) | **7b** |
| | Social influences on health perceptions | Any discussion or points in reference to the social influences on how participants perceive their health (either lung-related or not) from their social environment. For example, when participants relay information to SCPs about the incidence of lung problems in their social networks/community. Also code here when participants relay information to SCPs about the health damages of NRT products as perceived from media. | **7c** |
| | Accessibility to health resources | Any discussion or points in reference to participants' accessibility to health resources, i.e., accessibility to healthcare services or to health information. | **7d** |
| | Participants' motivation to quit | Any discussion or points that demonstrate participants' motivation to quit smoking and how it changed throughout the services. Also code here when SCPs reflect on why they think there were changes to motivation. | **7e** |
| | Participants' experiences of previous quit attempts prior to the study | Any discussion or points in reference to participant's previous smoking cessation attempts, the physical side-effects/emotional experiences participant experienced during these, and participant's attitudes towards behavioural support, NRT and e-cigarettes. Code here when SCP discuss participant's previous quit attempts prior to their participation in the study. For reflections on participants current quit attempts within the study, code at 9c. | **7f** |
| | Pre-existing health issues | Any discussion or points raised in reference to participant's pre-existing health issues (both physical and mental health). | **7g** |
| | Beliefs about smoking and smoking cessation | Any discussion or points in reference to SCP's perceptions of participant's internal drivers of smoking and quitting smoking, evaluative beliefs surrounding smoking and smoking cessation, including risk-minimising beliefs and self-efficacy, and/or cognitive dissonance of attitudes towards smoking (smoking as a way to cope with various difficult life stressors). | **7h** |
| | Wider context | Any additional discussion or points relating to the impact of wider context that do not fit within existing 'Contextual facilitators or barriers for participants to quit smoking' sub-themes. For example, discussion surrounding participants' finances (taking NRT products because they are expensive, quitting as cigarettes getting expensive, etc.) | **7i** |

| | | | |
|---|---|---|---|
| **Potential contamination**<br><br>*Consider intervention fidelity, exposure, and contamination* | | Any discussion or points in reference to instances/events/experiences that could cause/indicate study contamination (can be experienced by either SCPs or participants). Including but not exclusive to:<br>• additional smoking cessation or lung health information/guidance/support provided by a formal service or HCP for participants<br>• when participants' told SCPs about other public smoking and/or lung health campaigns<br>• when participants' told SCPs that they had contact with participants in the other study arm<br>• when SCP were told by participants' that they had discussed the consultations with other study participants<br>• when SCPs discuss instances and/or their experiences of delivering standard best practice and/or the intervention to more than one person at a time (e.g. a couple or a mother and daughter) or living in the same household<br>• instances of social diffusion (i.e. when SCPs were told by participants that they had discussed the study/intervention with people within their social network or community outside of the study)<br>• when SCPs discuss instances and/or experiences of delivering enhanced intervention (booklet + script) to a participant in control arm (spillover?) | **8** |
| **Other** | Acceptability of trial-specific procedures and data collection | Any discussion or points in reference to the acceptability of trial-related procedures (e.g., taking consent, filling questionnaire, explaining randomisation allocation, etc.), i.e., how they felt delivering trial-specific components and how these components influenced SCP's provided support. For acceptability of training about trial-specific procedures, code at 2c. | **9a** |
| | Acceptability of the Lung Health Check via YLST | Any discussion or points in reference to the acceptability of LHC-related components (e.g., LHC letter, YLST staff, etc.), i.e., how they felt about LHC-specific components and how these components influenced SCP's provided support. For how the LHC influenced quitting smoking, code at 7a. | **9b** |
| | Experiences quitting during the study | Any discussion or points about participant's attempts to quit smoking following first contact with the SCP on the van. This could be a quit attempt or reduction in smoking. For discussion about participant quitting prior to the study, code at 7f. For discussion about participant using NRT products (e.g., moving down the dosage), code at 6a. | **9c** |
| | Health consequences of stopping smoking | Any discussion or points in reference to health consequences of stopping smoking, i.e., SCP observed or was informed by participants any positive or negative changes in participant's health since stopping or reducing smoking. For example, SCP mentioned participant told them they had improved breathing, improved sense of taste/smell, etc. | **9d** |
| | Perceptions of other health-related services and tests (not smoking-relevant) | Any discussion or points in reference to other healthcare tests/examinations/procedures/services or healthcare workers and their possible impact the intervention/study. Code here when they discussed services that do not involve smoking cessation support. | **9e** |
| **Impact of COVID-19** | Impact of COVID-19 on SCP training | Any discussion or points in reference to their experiences with trial-related training as impacted by the COVID-19 pandemic and any associated alterations due to the pandemic. | **10a** |

| | | | |
|---|---|---|---|
| | Impact of COVID-19 on intervention acceptability | Any discussion or points in reference to the acceptability of the intervention as impacted by the COVID-19 pandemic and any associated alterations due to the pandemic, including proxy reflections from participants | **10b** |
| | Impact of COVID-19 on intervention comprehension | Any discussion or points in reference to participants' understanding of the intervention and their personal results as impacted by the COVID-19 pandemic and any associated alterations due to the pandemic, including proxy reflections from participants. Dual code at 10d if booklet delivery via telephone was considered to impact on comprehension | **10c** |
| | Impact of COVID-19 on influences of enhanced intervention | Any discussion or points in reference to the influences of the enhanced intervention (booklet + script) as impacted by the COVID-19 pandemic and any associated alterations due to the pandemic:<br>• Delivery/influences of personalized risk information from booklet<br>• Responses to risk information<br>• Self-efficacy<br>• Response-efficacy | **10d** |
| | Impact of COVID-19 on influences of standard best practice smoking cessation support | Any discussion or points in reference to the influences of standard best practice smoking cessation support as impacted by the COVID-19 pandemic and any associated alterations due to the pandemic:<br>• Influence of NRT products<br>• Influence of CO validation monitoring<br>• Contact with SCPs | **10e** |
| | Impact of COVID-19 on potential contamination | Any discussion or points in reference to instances/events/experiences that could cause/indicate study contamination (can be experienced by either SCPs or participants) as impacted by the COVID-19 pandemic and any associated alterations due to the pandemic. | **10f** |
| | Impact of COVID-19 on lung health check | Any discussion or points in reference to their perceptions of the lung health check as impacted by the COVID-19 pandemic and any associated alterations due to the pandemic. | **10g** |
| | Impact of COVID-19 on other trial-specific components | Any discussion or points in reference to their perceptions of other trial-specific components (e.g., taking consent, filling questionnaire, explaining randomisation allocation, etc.) as impacted by the COVID-19 pandemic and any associated alterations due to the pandemic. For example, the impact of PPE use on delivery of questionnaires. | **10h** |
| | Impact of COVID-19 on contextual factors | Any discussion or points in reference to how COVID-19 impacted SCPs or participant's life, health, beliefs, etc. (either SCP observed or was informed by participants) | **10i** |
| **Good quotes** | | Any quotes that are deemed as representative of an idea in a subtheme or a major theme, and could be included in publication | **11** |

### Supplementary material 8: trial participant interview coding framework (combined across timepoints)

| Major Theme | Sub Theme | Definition | Code No. |
|---|---|---|---|
| **Intervention usage**<br><br>*Consider intervention exposure and fidelity* | | Any discussion or points raised in reference to intervention delivery and subsequent experience of smoking cessation support from the SCP(s). i.e. what was said. Important to consider:<br>• barriers/enablers to intervention usage<br>• what participants have done with the booklet (e.g. put it on the fridge, thrown it away etc.) | **1** |
| **Intervention comprehension**<br><br>*Consider intervention exposure, fidelity and theory testing* | | Any discussion or points raised in reference to understanding the intervention i.e. the participants understanding their personal results. Important to consider when they are talking about their understanding and whether understanding is sustained:<br>• was it when the SCP was going through the booklet with them<br>• afterwards when reviewing it on their own<br>• on subsequent follow-up telephone calls with the SCP<br>• barriers/enablers to intervention comprehension (e.g. literacy issues) –<br>• any questions participants ask the interviewer here (e.g. sometimes around products or meanings of results/images in their booklet) | **2** |
| **Influence of intervention on theoretically grounded behavioural targets**<br><br>*Consider context, fidelity and theory testing* | | Any discussion or points raised in reference to the influence of the intervention on their knowledge, beliefs or behaviours associated with smoking, lung cancer, COPD, heart health, quit motivation and stopping smoking. Consider effects of different intervention components (personalised booklet, discussion with the SCP, co-location etc) on behavioural targets, and whether particular components are more effective. If no changes have been made – why not. | **3** |
| | Influence of NRT products | Any discussion or points raised in reference to the impact or influence that NRT products have had on their **current** smoking cessation/quit attempt as provided as part of the trial | **3a** |
| | Influence of CO validation monitoring | Any discussion or points raised in reference to the impact or influence that conducting CO validation monitoring has had on their current smoking cessation/quit attempt as part of the trial | **3b** |
| | Influence of personalised risk information booklet | Any discussion or points raised in reference to the impact/influence that the personalised booklet had had on their current smoking cessation/quit attempt as provided as part of the trial<br>• Images of their own CT scan results (lungs & heart)<br>• Immediate/short-term/long-term health benefits of stopping smoking | **3c** |
| | Influences of personalised risk | Any discussion or points raised in reference to the impact/influence that the personalised risk discussions with | **3d** |

| | | | |
|---|---|---|---|
| | information from SCP | the SCP had on their current smoking cessation/quit attempt as provided as part of the trial<br>• Enhanced delivery of behavioural support of risk information<br>• Concentration on enhanced self-and response efficacy | |
| | Responses to risk information (Low or no threat) | Any discussion or points raised that demonstrate a low or no threat response to risk information provided as part of the enhanced trial (intervention group) | **3e** |
| | Responses to risk information (High threat) | Any discussion or points raised that demonstrate a high threat response to risk information provided as part of the enhanced trial (intervention group). This can be 1) Low efficacy, defensive motivation, fear control (low intent to quit & negative attitudes/behaviours) OR 2) High efficacy, protection motivation, danger control (high intent to quit & positive attitudes/behaviours) | **3f** |
| | Self-efficacy | Any discussion or points raised that demonstrate previous, current or changes in personal self-efficacy i.e. an individual's belief in their ability/capability to carry out a behaviour/goal successfully. | **3g** |
| | Response-efficacy | Any discussion or points raised that demonstrate previous, current or changes in personal response-efficacy i.e. an individual's belief in whether the recommended behaviour (stopping smoking) will address the threat i.e. will stopping smoking improve their health or reduce health risks for them | **3h** |
| | Motivation and affect | Any discussion or points raised that demonstrate motivation and affect e.g. anxiety/reassurance from the above | **3i** |
| | Contact with SCP | Any discussion or points raised that demonstrate how frequency of contact/social support influenced behaviour change | **3j** |
| | Severity of disease | Any discussion or points raised that relate to the participants severity of disease or demonstration of how this could have affected their reaction/engagement with the enhanced intervention or stop smoking journey | **3k** |
| **Intervention acceptability**<br><br>*Consider exposure* | | Any discussion or points raised in reference to the acceptability of the intervention. Including but not exclusive to:<br>• how they felt being offered smoking cessation support at their lung health check appointment<br>• talking to the SCP/veiws on SCP role<br>• receiving standard SCI (details below) – try and pick apart components if possible<br>• receiving enhanced intervention components (details below) – try and pick components apart if possible<br>• smoking cessation support<br>• ongoing telephone support<br>• acceptability of co-location? | **4** |
| **Potential study contamination**<br><br>*Consider reach and contamination* | | Any discussion or points raised in reference to instances/events/experiences that could cause study contamination. Including but not exclusive to:<br>• additional smoking cessation or lung health information/guidance/support provided by a formal service or HCP | **5** |

| | | | |
|---|---|---|---|
| | | • public facing smoking and/or lung health campaigns<br>• contact with participants in the other study arm<br>• discussions with others about taking part<br>• instances of social diffusion | |
| **Other**<br><br>*Consider context* | Influence of the lung health check | Any discussion or points raised in reference to their experience and/or comprehension of the lung health check. Important to consider the starting point of the lung health check as a 'teachable moment'. | **6a** |
| | Experience of current or previous quit attempts/smoking reduction associated with the study | Any discussion or points raised in reference to their current or previous cessation journey following first contact with the SCP on the van. This could be a quit attempt or reduction in smoking | **6b** |
| | Health consequences of stopping smoking | Any discussion or points raised in reference to health consequences of stopping smoking i.e. have they reported any positive or negative changes in their health since stopping or reducing smoking | **6c** |
| | Other changes to 'health' made following lung health check/YESS intervention | Any discussion or points raised in reference to other health changes made following their lung health check or engagement in the YESS trial. i.e. increased physical activity or healthier eating habits or changes to taste and smell | **6d** |
| | Beliefs about smoking and smoking cessation | Any discussion or points raised in reference to internal drivers of smoking and quitting smoking. Evaluative beliefs surrounding smoking and smoking cessation, including risk-minimising beliefs and self-efficacy. Cognitive dissonance of attitudes towards smoking, smoking as a way to cope with various difficult life stressors. | **6e** |
| | Social influences on smoking and smoking cessation | Any discussion or points raised by the participant in reference to influences on smoking behaviour in the social environment, interpretation of the attitudes of other smokers | **6f** |
| | Previous quit attempts - Perceived effectiveness of stop smoking services and smoking cessation aids | Any discussion or points raised in reference to previous smoking cessation attempts and physical side-effects/emotional experiences during these. Attitudes towards behavioural support, NRT and e-cigarettes. Code here when they discuss previous quit attempts | **6g** |
| | Contextual barriers to stopping smoking | Any discussion or points raised in reference to contextual barriers reasons (e.g. age, pre-existing health conditions and competing priorities) in relation to lack of motivation or success in stopping smoking. Code here when they discuss this motivation to quit before attending the van | **6h** |
| | Wider context | Any discussion or points raised relating to the impact of wider context that do not fit within existing 'Other' sub-themes | **6i** |
| | Impact of smoking cessation on personal environment | Any discussion or points raised in reference to the impact of smoking cessation on personal environment i.e. since stopping smoking participants mention a reduction in smoke smell in their home/they have decided to make some changes to their home since noticing these changes. Includes comments made by themselves and when they report others having made comments. Also code changes to smell on their clothes. | **6j** |
| | Pre-existing health - Impact of existing relationship with | Any discussion or points raised in reference to their existing relationship with healthcare, experience of healthcare tests/examinations/procedures or healthcare workers and | **6k** |

| | | | |
|---|---|---|---|
| | healthcare or healthcare tests | their possible impact the intervention/study. This can include both positive and negative experiences/impact. Code here when they discuss their pre-existing health and comorbid conditions. | |
| | Perceptions of lung health | Any discussion or points raised in reference to social influences on perceptions of lung health from social network | **6l** |
| **Control participants** | Influence of NRT products | Any discussion or points raised in reference to the impact or influence that NRT products have had on their **current** smoking cessation/quit attempt as provided as part of the trial | **7a** |
| | Influence of CO validation monitoring | Any discussion or points raised in reference to the impact or influence that conducting CO validation monitoring has had on their current smoking cessation/quit attempt as part of the trial | **7b** |
| | Contact with SCP | Any discussion or points raised that demonstrate how frequency of contact/social support or contact with the SCP in general has influenced quit attempts | **7c** |
| | Acceptability of standard care for smoking cessation provided | Any discussion or points raised in reference to the acceptability of the standard care smoking cessation support provided. Including but not exclusive to:<br>• how they felt being offered smoking cessation support at their lung health check appointment<br>• talking to the SCP/veiws on SCP role<br>• smoking cessation support<br>• ongoing telephone support<br>• acceptability of co-location? | **7d** |
| **Impact of COVID-19** | Impact of COVID-19 on lung health check | Any discussion or points raised in reference to their experience and/or comprehension of the lung health check as impacted by the COVID-19 pandemic and any associated alterations due to the pandemic | **8a** |
| | Impact of COVID-19 on intervention usage | Any discussion or points raised in reference to intervention delivery and subsequent experience of smoking cessation support from the SCP(s) that has been impacted by the COVID-19 pandemic and any associated alterations due to the pandemic | **8b** |
| | Impact of COVID-19 on intervention comprehension | Any discussion or points raised in reference to understanding the intervention i.e. the participants understanding their personal results as affected by the COVID-19 pandemic and any associated alterations due to the pandemic | **8c** |
| | Impact of COVID-19 on the influence of intervention on theoretically grounded behavioural targets | Any discussions or points raised that infer an impact of COVID-19 on any of the below (as expanded above):<br>• Influence of NRT products<br>• Influence of CO validation monitoring<br>• Influence of personalised risk information booklet<br>• Influences of personalised risk information from SCP<br>• Responses to risk information (Low or no threat)<br>• Responses to risk information (High threat)<br>• Self-efficacy<br>• Response-efficacy<br>• Motivation and affect<br>• Contact with SCP | **8d** |

| | | | |
|---|---|---|---|
| | | • Severity of disease | |
| | Impact of COVID-19 on intervention acceptability | Any discussion or points raised in reference to the acceptability of the intervention as impacted by the COVID-19 pandemic and any associated alterations due to the pandemic | **8e** |
| | Impact of COVID-19 on potential study contamination | Any discussion or points raised in reference to instances/events/experiences that could cause study contamination as impacted by the COVID-19 pandemic and any associated alterations due to the pandemic | **8f** |
| | Contextual impact of COVID-19 | Any discussion about how covid-19 has impacted the participant's life, health etc. | **8g** |
| | Impact of COVID-19 on acceptability and delivery of standard smoking cessation support | Any discussion or points made raised relating to the acceptability and delivery of standard smoking cessation support during the pandemic i.e. altered procedures where needed (generally relevant to the control group) | **8h** |
